# AI Agents in Clinical Medicine: A Systematic Review

**DOI:** 10.1101/2025.08.22.25334232

**Authors:** Alon Gorenshtein, Mahmud Omar, Benjamin S Glicksberg, Girish N Nadkarni, Eyal Klang

## Abstract

**Background:** AI agents built on large language models (LLMs) can plan tasks, use external tools, and coordinate with other agents. Unlike standard LLMs, agents can execute multi-step processes, access real-time clinical information, and integrate multiple data sources. There has been interest in using such agents for clinical and administrative tasks, however, there is limited knowledge on their performance and whether multi-agent systems function better than a single agent for healthcare tasks.

**Purpose:** To evaluate the performance of AI agents in healthcare, compare AI agent systems vs. standard LLMs and catalog the tools used for task completion

**Data Sources:** PubMed, Web of Science, and Scopus from October 1, 2022, through August 5, 2025.

**Study Selection:** Peer-reviewed studies implementing AI agents for clinical tasks with quantitative performance comparisons.

**Data Extraction:** Two reviewers (A.G., M.O.) independently extracted data on architectures, performance metrics, and clinical applications. Discrepancies were resolved by discussion, with a third reviewer (E.K.) consulted when consensus could not be reached.

**Data Synthesis:** Twenty studies met inclusion criteria. Across studies, all agent systems outperformed their baseline LLMs in accuracy performance. Improvements ranged from small gains to increases of over 60 percentage points, with a median improvement of 53 percentage points in single-agent tool-calling studies. These systems were particularly effective for discrete tasks such as medication dosing and evidence retrieval. Multi-agent systems showed optimal performance with up to 5 agents, and their effectiveness was particularly pronounced when dealing with highly complex tasks. The highest performance boost occurred when the complexity of the AI agent framework aligned with that of the task.

**Limitations:** Heterogeneous outcomes precluded quantitative meta-analysis. Several studies relied on synthetic data, limiting generalizability.

**Conclusions:** AI agents consistently improve clinical task performance of Base-LLMs when architecture matches task complexity. Our analysis indicates a step-change over base-LLMs, with AI agents opening previously inaccessible domains. Future efforts should be based on prospective, multi-center trials using real-world data to determine safety, task matched and cost-effectiveness.

**Primary Funding Source:** This work was supported in part through the computational and data resources and staff expertise provided by Scientific Computing and Data at the Icahn School of Medicine at Mount Sinai and supported by the Clinical and Translational Science Awards (CTSA) grant UL1TR004419 from the National Center for Advancing Translational Sciences. Research reported in this publication was also supported by the Office of Research Infrastructure of the National Institutes of Health under award number S10OD026880 and S10OD030463. The content is solely the responsibility of the authors and does not necessarily represent the official views of the National Institutes of Health.

**Registration:** PROSPERO CRD420251120318

## INTRODUCTION

Large language models (LLMs) have demonstrated expert performance on medical examinations^1,2,3^. Recent studies have begun to implement these LLMs in clinical settings, showing improvements in performance.^4,5^ However, clinical deployment faces critical safety challenges^6,7,8,9^ LLMs can generate plausible but incorrect information, known as hallucinations.^10,11^ In up to 22% of clinical interactions, LLM showed misinformation errors most common in medication dosing, symptom recognition, and diagnostic reasoning.^12,13^ These models also may lack access to current medical literature and patient-specific data from electronic health records, limiting their ability to provide evidence-based care.^14,15^

To overcome these limitations, researchers have developed AI agents, LLM systems augmented with tools that can access external information, perform calculations, and complete multi-step clinical tasks autonomously.^16–20,21,22^ Unlike standard LLMs that provide single responses to questions, AI agents can search medical databases, extract data from patient records, calculate medication doses, and coordinate multiple information sources to answer complex clinical questions.^23,24,25^

Multi-agent systems (Agentic AI)^26^ take this further, using teams of specialized AI agents^27^ with an orchestration layer that collaborate like multidisciplinary care teams to tackle complex interdisciplinary diagnostic and treatment planning challenges.^28,29,30,31^ Despite their theoretical advantages over base LLMs, systematic evidence on the use of AI agents in clinical tasks is lacking.

This systematic review evaluates the current state of AI agents in clinical medicine, quantifies their performance compared with base LLMs, and identifies applications suited for research and near-term integration. We also outline safeguards and design principles needed to ensure safe and effective implementation.

## METHODS

### Protocol and Registration

This study used an established methodological framework, consistent with the extended Preferred Reporting Items for Systematic Reviews and Meta-Analyses (PRISMA) checklist for systematic reviews.^32^ The study protocol was pre-registered on the International Prospective Register Reviews (PROSPERO; CRD420251120318).

### Data Sources and Searches

We conducted a comprehensive literature search across three bibliographic databases: PubMed, Web of science, and Scopus, from October 1, 2022 (corresponding to the rise of agentic frameworks) through August 5, 2025. The search strategy combined Medical Subject Headings (MeSH) terms and free-text keywords across three conceptual domains: (1) agentic systems ("multi-agent" OR "tool-calling" OR "agentic AI" OR "autonomous agent" OR "ReAct" OR "LangChain" OR "AutoGen"); (2) large language models ("GPT" OR "Claude" OR "Gemini" OR "LLaMA" OR "large language model" OR "foundation model"); and (3) clinical applications ("clinical" OR "medical" OR "healthcare" OR "diagnosis" OR "treatment" OR "patient care"). The full search strategy for each database is provided in Supplementary Methods. We additionally performed forward and backward citation searching of included studies and reviewed preprint servers (arXiv, medRxiv, bioRxiv) to identify articles in press. For articles that have not yet undergone peer review, we present a narrative analysis outside the rigorous systematic review criteria.

### Study Selection

Studies were included if they met all of the following criteria: (1) described an AI agent system, defined as an LLM that autonomously determines when to call external tools, coordinates multiple specialized agents, or both; (2) applied this system to a human health-related task with clinical relevance; (3) reported quantitative performance metrics comparing the agentic approach to a baseline (single-pass LLM, human performance, or established clinical standard); (4) were published as peer-reviewed articles in English.

We excluded: (1) studies using single-pass LLMs without agentic capabilities; (2) purely technical papers without clinical applications; (3) simulation or benchmark studies lacking real-world validation or clinical tasks; (4) review articles, editorials, conference abstracts, and preprints not yet accepted for publication.

Two reviewers (A.G, M.O) independently screened all titles and abstracts using Covidence systematic review software. Articles marked for potential inclusion by either reviewer underwent full-text assessment. Disagreements were resolved through discussion, with a third reviewer (E.K) providing arbitration when consensus could not be reached.

### Data Extraction and Quality Assessment

We developed a standardized data extraction form, piloted on five randomly selected studies, and refined based on extractor feedback. Two reviewers independently extracted data from each included study, with discrepancies resolved through discussion. Extracted variables included:

1. Study characteristics: First author, publication year, country, clinical domain, study design, sample size, data source (real-world vs. synthetic), external validation (yes/no), code availability.

Technical specifications: Base-LLM(s) used, AI agent framework (e.g., AutoGen, LangChain, CrewAI and custom)^33^, agent architecture type (multi-agent only, tool-calling only, or combined), number of agents, number and types of tools, and consensus mechanism for multi-agent systems.

Performance metrics: Primary outcome measure, baseline comparator, absolute and relative performance improvement, secondary outcomes, and subgroup analyses where available.

Implementation details: Computational requirements, runtime, and cost estimates were reported, and human oversight requirements were reported.

### Risk of Bias Assessment

We adapted the QUADAS-AI tool^34^ (Quality Assessment of Diagnostic Accuracy Studies for AI) to assess risk of bias and applicability concerns. This modified instrument evaluated four domains: (1) Patient/Data Selection, (2) Index Test (AI system), (3) Reference Standard, and (4) Flow and Timing. Each domain was rated as low risk, high risk, or unclear risk of bias.^34^ Additionally, we assessed concerns regarding applicability for the first three domains.

Specific high-risk indicators included: use of synthetic/simulated data only, single-center validation, sample size <100, lack of external validation, unclear ground truth establishment, and selective outcome reporting. Two reviewers independently assessed each study, with disagreements resolved through consensus discussion.

### Data Synthesis and Analysis

Given the heterogeneity in clinical domains, outcomes, and architectures, we did not conduct a meta-analysis; instead, we performed a narrative synthesis structured around our pre-specified research questions. We stratified analyses by:

1. Architecture type: Studies were categorized as single AI agent using tool-calling only, multi-agent only, or combined (multi-agent plus tools) based on their primary implementation.
2. Clinical domain: We grouped studies into diagnosis/prognosis, evidence synthesis, treatment planning, clinical operations, genomics, and other specialized applications.
3. Complexity metrics: We analyzed relationships between performance and system complexity, defined by number of agents (for multi-agent systems) and number of tools (for tool-calling systems).

For studies reporting comparable outcomes, we calculated median performance improvements with interquartile ranges (IQR).

## Results

### Study Selection

Our search strategy across PubMed, Web of Science, and Scopus identified 6,023 records after removal of duplicates. Following title and abstract screening, 254 articles underwent full-text assessment for eligibility. After applying inclusion criteria requiring peer-reviewed AI agent LLM implementations for clinical tasks, 20 studies met all criteria and were included in the final analysis.^35–53,54^ Reasons for exclusion included: non-AI agent single-pass LLM implementations (n=184), simulation or benchmark-only studies without clinical validation (n=31), and studies lacking comparative performance metrics (n=22) (**Supplementary Figure 1.**).

### Overview of the included studies

The 20 included studies, published between 2024-2025, encompassed many clinical applications with different evaluation datasets: clinical cases (n=5, ranging from 16-302 cases), clinical reports/EMG interpretations (n=2, totaling 419 reports), multiple-choice questions (n=2, totaling 5,120 MCQs), evidence synthesis queries (n=4, ranging from 50-500 queries), actual patient data (n=2, 117 patients), computational vignettes (n=1, 10,000 calculations), biological/genomic datasets (n=4, including 8 biomarker datasets, 92 nanobodies, 1,106 gene sets, and 272 articles) (**Supplementary Table 1**). Sample sizes varied from 8 to 10,000 items. Studies were predominantly comparative designs (n=16, 80%), with two simulation studies validated through prospective laboratory experiments (10%), one retrospective case-control study (5%), and one randomized controlled trial (5%). Clinical domains clustered around decision-support tasks: diagnosis and prognosis applications dominated (n=8, 40%), particularly for rare disease diagnosis, followed by evidence synthesis (n=5, 25%), treatment planning (n=3, 15%), clinical operations (n=2, 10%), genomics applications (n=2, 10%), and medical education (n=1, 5%). Agent architectures revealed three implementation patterns: single-agent tool-calling frameworks (n=8, 40%), multi-agent systems without tool integration (n=5, 25%), and hybrid multi-agent plus tool-calling architectures (n=7, 35%). The predominant LLM backbone was the GPT-4 family (n=15, 75%), followed by Llama- 3 variants (n=4, 20%), Claude-3 Opus (n=4, 20%), and Gemini-1.5 (n=4, 20%), with multiple studies testing several models. Notably, only one study employed prospective randomized controlled trial methodology.

### Risk of Bias

Quality assessment using QUADAS-AI criteria revealed some methodological concerns. Only 30% (6/20) of studies achieved low risk of bias across all domains. The remaining 70% demonstrated high risk in at least one domain, with patient selection representing the most problematic area (60% high risk). Other sources of bias stemmed from: synthetic data dependence (n=13, 65%), single-center designs (n=20, 100%), spectrum bias, reference standard concerns (n=5, 25%) (**Supplementary Table 1**).

### Performance Synthesis

#### Overall Performance Improvements

Aggregate performance improvements demonstrated substantial heterogeneity, with median gains of +36% over baseline LLM (range: +3.5% to +76%).

#### Single AI Agent Tool-Calling Performance

Single-agent implementations were typically deployed for narrow clinical domains to address specific LLM limitations. These architectures achieved the highest median improvement (+53.0%, IQR: 36.0-56.9%) (**Table 2**). Our analysis identifies two primary reasons for this performance increase: one focuses on enhancement through tools (independent of the AI agent framework), while the other pertains to the AI agent framework itself. Goodell et al., Xu et al., and Low et al. primarily achieved performance gains through the tools they employed. Goodell et al.’s calculation tool, which is crucial for LLMs given their inherent challenges with computations^55^ improved performance by +59.1% in comparison to the base-LLM (Gpt-4o). Woo et al., Low et al., and Xu et al. showed improved evidence gathering using domain-specific web search for oncology, orthopedics, and genomics. Web search in the studies we found was the most common tool implementation (n=8). In contrast, Ferber et al. observed that clinical decision-making in oncology necessitates capabilities beyond standard LLMs and tools, including multi-modal interpretation of pathological biopsies, radiographic imaging, and complex calculations, which led to the development of their AI agent. The AI agent surpassed baseline LLM (GPT-4) performance by 56.9%. Similarly, Pickard et al. developed an AI agent for biomarker discovery, yielding improved results compared to both the base-LLM and tool augmentation LLM.

**Table 1.**
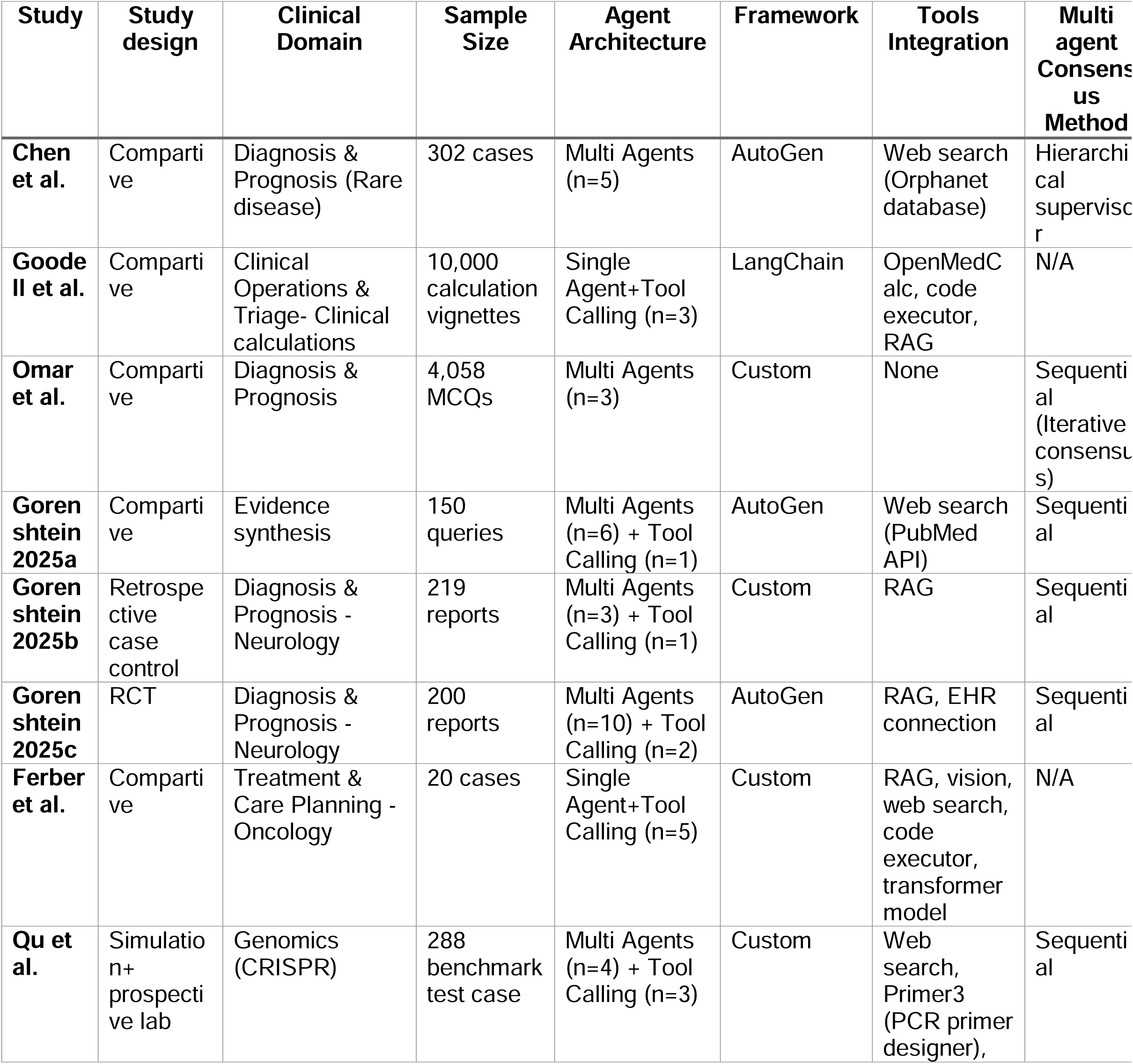
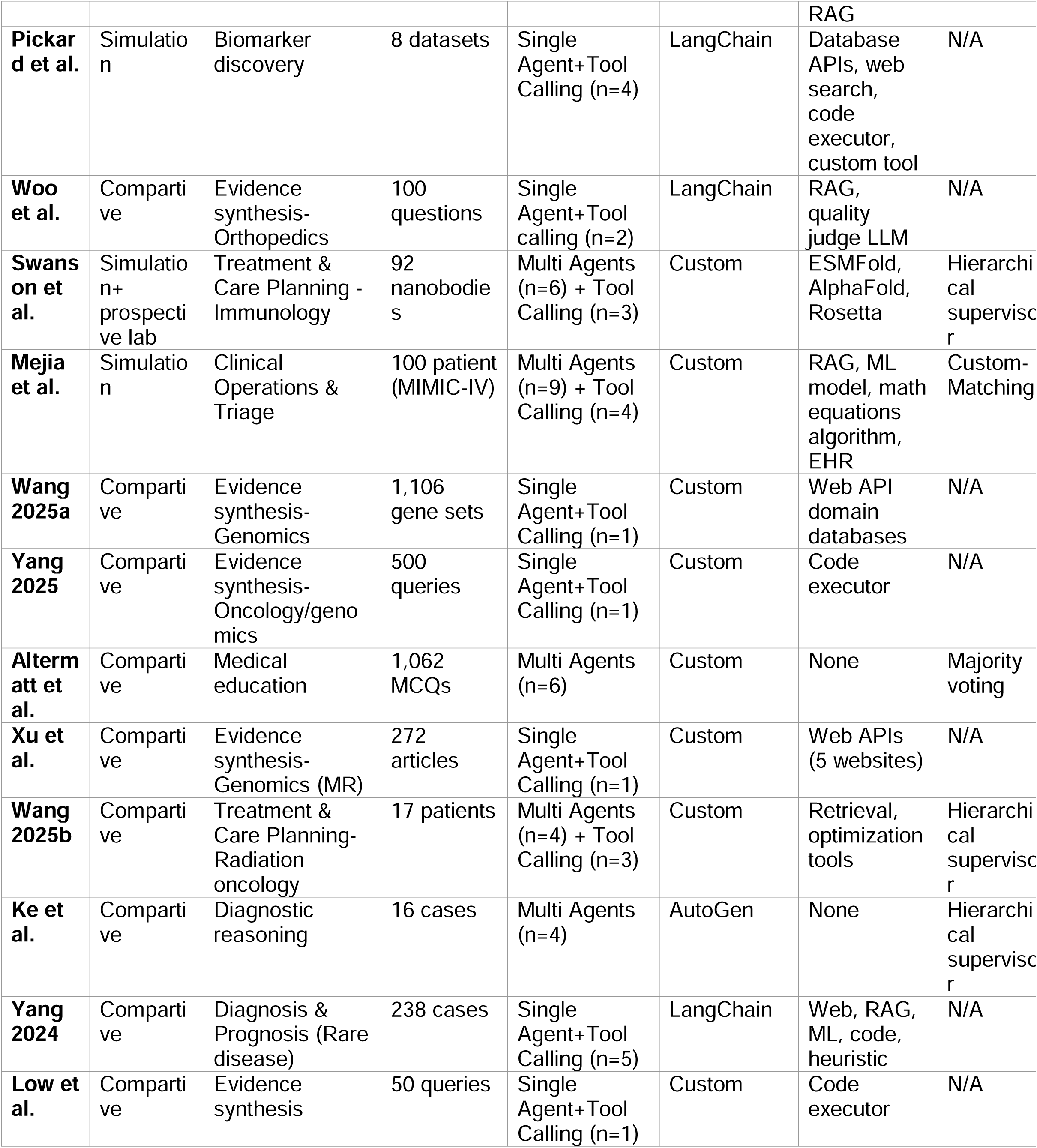
Study Characteristics.

**Table 2.** Clinical performance of agent versus baseline.

| **Study** | **AI Agent Base-LLM Model(s)\*** | **Primary Outcome** | **Agent vs Baseline\*** | **Baseline comparison** | **External validation** | **Synthetic Data** |
| --- | --- | --- | --- | --- | --- | --- |
| **Chen et al.** | GPT-4 | Accuracy | +14.3% | Base-LLM | No | Yes |
| **Goodell et al.** | GPT-4o, Llama-3 | Accuracy | +59.1% | Base-LLM | Yes | Yes |
| **Omar et al.** | GPT-4o, Claude, Gemini-1.5-Pro | Accuracy | +13.8% | Base-LLM | No | No |
| **Gorenshtein 2025a** | GPT-4o-mini | Accuracy | +3.5% | Single LLM (Sonar-pro) | No | Yes |
| **Gorenshtein 2025b** | Gemini-1.5-Pro-002 | Accuracy | +29.6% | Base-LLM | Yes-Physician review | No |
| **Gorenshtein 2025c** | GPT-4o | Quality score | -25.5% | Physicians | Yes-Comparison to Physician | No |
| **Ferber et al.** | GPT-4 | Accuracy | +56.9% | Base-LLM | Yes-Physician review | Mixed |
| **Qu et al.** | GPT-4o, CRISPR-Llama3-8B (fine tuned Llama3-8b) | Design success | +49% | Base-LLM | Yes- in-vitro validation | Mixed |
| **Pickard et al.** | GPT-4 | Enrichment similarity score | +61.7% | Base-LLM | Yes | No |
| **Woo et al.** | GPT-4, Claude-3, Llama-3-70B, GPT-3.5, Mistral-8x7B | Accuracy | +36% | Base-LLM | Yes-Physician review | Yes |
| **Swanson et al.** | GPT-4o | Binding improvement | 2/92 improved | Baseline wild-type nanobodies (no agent design) | Yes- in-vitro validation | Mixed |
| **Mejia et al.** | GPT-4o, GPT-4o-mini, Qwen-2.5-32B, DeepSeek-R1 | Custom score | N/A | Base-LLM | No | No |
| **Wang 2025a** | GPT-4 | ROUGE-L | +7.1% | Base-LLM | No | No |
| **Yang 2025** | Llama-3 | None | N/A | None | No | Yes |
| **Altermatt et al.** | GPT-4o | Accuracy | +4.1% | Base-LLM+chain of thoughts | No | Yes |
| **Xu et al.** | GPT-4-Turbo, GPT-3.5-Turbo, Mixtral- | Accuracy | N/A | None | Yes | Yes |
|  | 8x22B, Llama-3 8B, Llama-3 70B, Qwen 2.5-max, Claude-3-Opus |  |  |  |  |  |
| **Wang 2025b** | GPT-4o | PTV D95 | +4.75% | ECHO auto-planner | Yes | Yes |
| **Ke et al.** | GPT-4-Turbo | Accuracy | +76% (Base-LLM) 28% (Physician) | Base-LLM, Physicians | Yes-Comparison to Physician | Yes |
| **Yang 2024** | GPT-3.5-Turbo | F1 score | +53% | Base-LLM | Yes | Yes |
| **Low et al.** | ChatRWD | Accuracy | +48% | Chatgpt-4 | No | Yes |

\*The comparison to Baseline unless described otherwise is to Base-LLM. This was conducted on the best performance agentic LLM backbone (e.g. ChatGPT-4 Agentic compared to Chatgpt-4). Percentage point difference

#### Multi Agents performance

Our systematic review identified two distinct multi-agent approaches. Pure multi-agent systems without tools showed modest gains (+14.05%, IQR: 8.95-45.15%). Multi-agent systems with tools demonstrated slightly better performance (+17.17%, IQR: 4.12-39.3%) with high variance (**Table 2**). The reason for the high variance may be as these multi-agent frameworks were often applied to tasks that could have been efficiently managed by either a single agent or tool-augmented LLMs (non-AI agent framework). Nevertheless, for the studies who showed high increased performance was particularly noteworthy in more complex tasks, indicating that multi-agent systems may be more beneficial in such scenarios.

Qu et al. developed a multi-agent team incorporating a fine-tuned CRISPR-Llama3 model, completing 22 gene-editing tasks on 288 benchmark test cases and achieving external validation through AI-guided knockout of four genes (TGFβR1, SNAI1, BAX, BCL2L1). Swanson et al. created a "virtual laboratory" comprising specialized agents (PI, immunologist, machine learning specialist, etc.), successfully developing antibodies validated through wet-lab experiments. Wang 2025 developed a multi-agent system for oncology treatment planning, achieving +4.75% improvement over standard ECHO auto-planning methods for lung cancer. Moreover, Ke et al. demonstrated that multi-agent systems can mitigate clinical decision biases, improving accuracy from 0% to 76% on bias-containing complex cases, surpassing human physicians by 21%. Chen et al. similarly showed that multi-agent improved reasoning for rare disease diagnosis.

Mejia et al. demonstrated managing separately different tools by advanced custom multi-agent frameworks that can automate appointment scheduling by efficiently matching patients with resources. However, the benefits of multi-agent systems tend to be limited for tasks that are already suited for single AI agents. Gorenshtein 2025a, Chen et al., and Altermatt et al., who utilized multi-agent systems for evidence synthesis and answering multiple-choice questions, showed that the improvements are primarily linked to the tools implemented rather than the collaborative efforts of the agents. As mentioned by Altermatt et al. that simpler single-agent strategies are sufficient to address most of the questions in the study.^48^

#### Agent Number and Tool Scaling Effects

Our analysis revealed an inverted-U relationship between agent number and performance, with optimal results at 4-5 agents before declining performance (β = - 8.815, R² = 0.162). Tool number showed a weak positive correlation with performance (β = 8.869, R² = 0.377). It’s important to consider that our analysis is highly limited by task heterogeneity and varying comparison baselines (**Supplementary Figure 2**).

#### Multi agent Consensus Method

Multi-agent implementations employed various consensus mechanisms: supervisor agent coordination (36.36%, n=4/11), sequential processing (45.45%, n=5/11), majority voting (9.09%, n=1/11), and custom approaches (9.09%, n=1/11).

#### Base-LLM vs AI agent deployment and backbone suitability

We analyzed models with paired baselines and AI agent variants to determine which LLM might be most beneficial for implementing AI agent workflows. We did not find any specific LLM that would benefit significantly more from the AI agent workflow. However, we observed that older models with lower performance might have greater improvement potential.

When inspecting backbone suitability for an AI agent framework, Ferber et al. noted that smaller parameter models, such as Llama-3-70B and Mixtral-8×7B, struggle to provide accurate timing for tool calling compared to higher preforming models like GPT-4 (87.5% for GPT-4 vs. 39.1% for Llama and 7.8% for Mixtral). Similarly, Goodell et al. demonstrated lower performance from Llama-3-70B compared to GPT-4 as backbones of AI agent systems. In examining larger parameter models, Omar et al. did not observe any significant differences regarding the backbone of the LLM itself. Conversely, Woo et al. and Mejia presented conflicting findings; however, Woo et al. study was simple enough that the tool itself was enough to solve it, and Meija study utilized different outcomes, making it challenging to relate their results to this comparison (**Figure 1**.).

**Figure 1.**
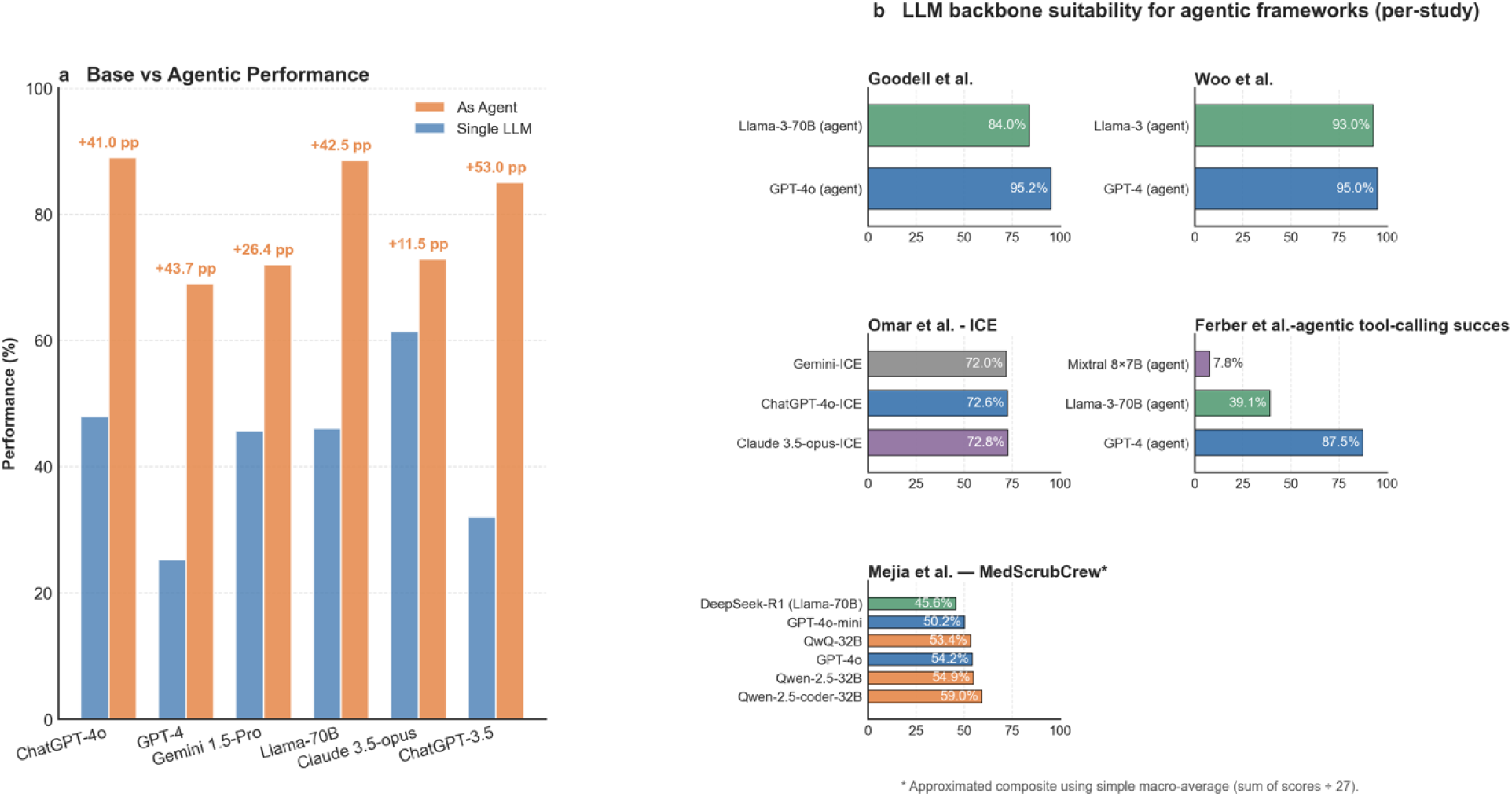
Agentic LLM backbones for clinical tasks: uplift over single-model baselines and per-study comparisons a, “Base vs agentic” performance for five backbones. Bars show accuracy/success (%) reported in the cited sources; orange = *As agent*, blue = *Single LLM*. Numbers above bars are percentage-point (pp) uplift. Values: ChatGPT-4o 88.93 vs 47.91 (+41.0 pp); GPT-4 68.97 vs 25.23 (+43.7 pp); Gemini-1.5-Pro 71.97 vs 45.58 (+26.4 pp); Llama-3-70B 88.50 vs 46.00 (+42.5 pp); Claude-3.5-opus 72.83 vs 61.34 (+11.5 pp). b, Per-study agentic comparisons. Goodell et al. (clinical calculations): GPT-4o 95.2, Llama-3-70B 84.0. Woo et al. (orthopedic guideline QA): GPT-4 95.0, Llama-3 93.0. Omar et al. - ICE (final agentic versions): Claude-3.5-opus-ICE 72.83, ChatGPT-4o-ICE 72.59, Gemini-ICE 71.97. Ferber et al.: GPT-4 87.5, Llama-3-70B 39.1, Mixtral-8×7B 7.8. Mejia et al. - MedScrubCrew* (overall composite): Qwen-2.5-coder-32B 59.0; Qwen-2.5-32B 54.9; QwQ-32B 53.4; GPT-4o 54.2; GPT-4o-mini 50.2; DeepSeek-R1 (Llama-70B) 45.6. *\**Asterisk denotes an approximated composite (macro-average of 27 judged items).

### Preprint AI agents’ frameworks

Notable preprint studies not included in the formal analysis demonstrated advancing capabilities of multi-agent frameworks. ^24,30,56,57^ The Microsoft MAI-DxO framework achieved 85.5% diagnostic accuracy while reducing costs by 69% compared to baseline models (from $7,850 to $2,397 for comparable accuracy). ^21^ Li et al. reported progressive performance improvements in a simulated hospital environment across 32 departments, with agents learning from sequential patient interactions. ^49^ Integration with electronic health records was demonstrated by Shi et al., showing 29.6% improvement over baseline in multi-tabular reasoning tasks.^48^ For radiological applications, Chen et al. implemented a modular multi-agent system achieving 79.9% overall diagnostic accuracy in chest X-ray interpretation (**Supplementary Table 2**).^58^

## Discussion

This systematic review demonstrates that AI agents consistently improve clinical task performance when architecture matches task complexity. Shown by the heterogeneity in performance improvements, ranging from +3.5% to +76%, with greater enhancements and narrow range observed when the architecture corresponded to task complexity.

Our analysis indicates a step-change over base-LLMs, with AI agents opening previously inaccessible domains **(Figure 2**.).

**Figure 2.**
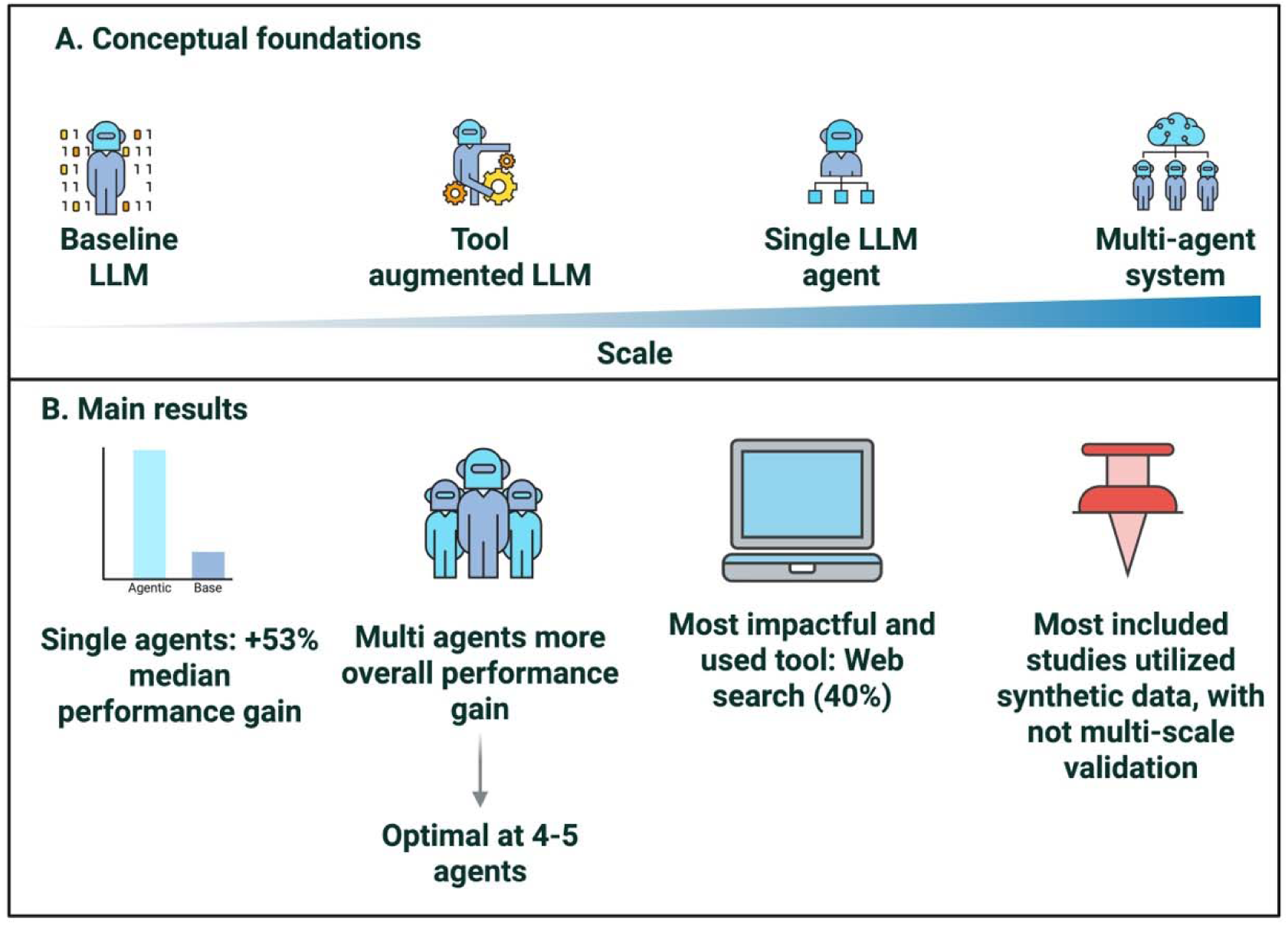
Conceptual foundations of LLM agent systems and a summary of the main results of the review. **A.** The top panel shows the progression in agentic scale from a baseline LLM to a tool-augmented LLM, a single LLM agent, and a multi-agent system. **B.** The bottom panel summarizes findings of the review: single agents produced a median 53% performance gain over base LLMs; multi-agent systems delivered larger benefits for complex tasks with optimal teams of 4–5 agents; web search was the most commonly used and most impactful tool (about 40% of studies); many studies relied on synthetic data and lacked multi-site validation. The gradient indicates increasing agentic complexity and coordination.

### Defining AI agents in Healthcare

The inconsistent use of "AI agent" terminology in medical literature^59,60^ necessitates clear definitions. Based on the literature^16–20,27^ and our analysis, we identified the following definitions. A true AI agent requires a minimum two of three core capabilities with the first core being mandatory, while for Agentic AI (multi-agent) both the first and the third core are mandatory: 1-Iterative reasoning processes (e.g., ReACT prompting^61^)-mandatory, 2-Tool selection, 3-Multi-agent collaboration. By adopting this terminology, we believe future clinical studies can be more accurately categorized into LLM, LLM utilizing tools, AI agents, and multi-agent systems (Agentic AI) (**Figure 2**.).

### Practical Considerations for Implementation

Our analysis reveals several critical factors that institutions should consider when implementing AI agent systems in clinical practice.

### Choosing the Right Model Architecture

While no single LLM emerged as universally superior for AI agent applications, model size significantly impacts performance. Lower performance models struggled with the complex reasoning required for tool selection. For example, Ferber et al. found that GPT-4 achieved 87.5% accuracy in tool-calling decisions compared to only 39.1% for Llama-3-70B and 7.8% for Mixtral-8B. This finding contradicts recent enthusiasm for small language models (SLMs)^62^ in healthcare and suggests institutions should prioritize higher performance, well-validated models for initial implementations.

However, a promising hybrid approach may reconcile cost concerns with performance needs: using a high performance "orchestrator" model to interpret findings and coordinate tasks (deciding which tool and which agent to use), while deploying smaller, specialized models (SLMs) as "workers" for specific functions (using the tool themselves without thinking).^63^ In this architecture, the orchestrator handles complex reasoning and tool calling, while SLMs execute defined tasks like data extraction or calculation. This approach could reduce computational costs while maintaining the sophisticated reasoning necessary for clinical decision-making.

### Optimal Team Size for Multi-Agent Systems

For institutions considering multi-agent implementations, our analysis identified a highest performance for team of 4-5 agents, beyond which performance plateaus or even declines (**Supplementary Figure 2A**). This finding aligns with established organizational theory on optimal team sizes^64^ and challenges the assumption that adding more agents inevitably improves outcomes.^65^

### Tool Calling and Integration

The number of tools showed a modest positive correlation with performance (Figure 2B), with web search emerging as the most consistently valuable addition (40% of the studies). For clinical deployment, we found that most framework have the following core set of validated tools: medical literature search (RAG or web-search domains e.g. PubMed) for evidence-based recommendations, calculator functions for dosing and risk scores if needed, structured data extraction from clinical notes and current clinical guideline access. Additional tools can be added incrementally based on specific workflow needs rather than implemented all at once.

### Consensus Mechanisms: Matching Method to Task

When implementing multi-agent systems, the choice of consensus mechanism matters. Our analysis identified three primary approaches with different strengths: Supervisor coordination (36.4% of studies): Best for hierarchical clinical decisions where one agent needs final authority, Sequential processing (45.5%): Optimal for stepwise workflows like diagnostic workups, Democratic voting (9.1%): Most effective for consensus-building in ambiguous cases. Recent evidence suggests task-specific optimization: voting protocols excel in diagnostic reasoning tasks (13.2% improvement over consensus) by exploring multiple diagnostic pathways, while consensus methods perform better for factual queries (+2.8%) through cross-verification.^66^ Institutions should match the consensus mechanism to their specific use case rather than defaulting to one approach.

### Implementation Recommendations

Based on our findings, we provide a structured approach to implementing AI agents. Initially, it is advisable to utilize high-performing models as the primary reasoning model, acting as the “orchestrator.” Additionally, it is important to limit the use of multi-agent systems to no more than five specialized agents to maintain clarity and efficiency.

Institutions should start with essential tools, such as search functionalities, calculations, and data extraction capabilities and expand their toolkit as demonstrated needs arise. Furthermore, aligning consensus mechanisms with clinical workflows is crucial, as a tailored approach will be more effective than a one-size-fits-all solution. Lastly, considering hybrid architectures that strike a balance between sophisticated reasoning and computational efficiency, such as employing “workers” and SLM for tool-using agents, can enhance overall performance. These practical considerations, derived from our systematic analysis, provide early roadmap that will require further research to validate this approach. Institutions and researchers following this roadmap may begin their journey with AI agents while avoiding common pitfalls as identified in early implementations studies.

### Architecture-Task Alignment

In our analysis, we observed a pattern indicating that more complex frameworks tend to yield better performance for intricate tasks. Based on this finding and clinical readiness indicators, we built upon the results a practical three-tier framework for clinicians considering the implementation of AI agents. These tiers based on three criteria: (1) consistency of performance improvement across studies, (2) quality of validation data (real-world vs. synthetic), and (3) implementation complexity and risk. This framework will help clinicians avoid both under-utilization of beneficial tools and over-engineering of simple problems.

#### Tier 1 - Ready for Clinical Pilot Programs

Tool-augmented LLMs for discrete, auditable micro-tasks (medication calculations, Hounsfield unit extraction).^67,68^ These showed +53% median improvement and address specific limitations of standard LLMs, with low bias in the way the tool is being implemented. They primarily rely on Base-LLM without significant modifications, suggesting a safe application in clinical practice.^69,70^ Woo et al. illustrated that sometimes, the solution lies simply in utilizing tool-augmented LLMs. They compared LLMs integrated with RAG to a single AI agent utilizing RAG (Tier-2), revealing only a 2% performance increase (+34% compared to the baseline of ChatGPT-4) when responding to questions concerning orthopedic guidelines.

#### Tier 2 - Supervised Clinical Research

Single AI agents are well suited to manage complete clinical workflows (e.g., report generation, treatment recommendations, differential diagnosis development)^58,71^. The single AI agent show promise but require careful human oversight. These systems achieved strong performance gains (Ferber et al. +57%, Gao et al. +17%) but rely heavily on synthetic data and lack real-world validation. We recommend these be tested in supervised research settings with "human-in-the-loop" mechanisms, allowing clinicians to review and modify outputs before implementation.^72,67^ This is particularly important given the ethical considerations of autonomous clinical decision-making.^75,76^ Instead, clinicians should explore, using real clinical data, how iterative reasoning processes can enhance the performance of tool-augmented LLMs in tasks that demand more than simple tool usage

#### Tier 3 - Research Only

Multi-agent systems, despite theoretical appeal, showed high variability, modest improvements over single agents, and in one randomized trial, performed worse than physicians. Our analysis found diminishing returns beyond 4-5 agents and suggests these systems are often over-engineered for problems solvable with simpler approaches. Reserve these for genuine interdisciplinary challenges in controlled research environments.

This hierarchical framework function as a three-layer framework where the clinicians should always try implement their choice of model from the lower tier and move to the next tier if the results aren’t sufficient (**Figure 3**.). By this logic the physicians will benefit by having a well fitted model, avoiding unnecessary computational cost or high amount of errors. Additionally, researchers will benefit from knowing at which clinical stage they should implement the three framework provided (tool-augmented LLMs, Single AI agent, Multi AI agents) (**Table 3**, **Supplementary table 3,4**).

**Figure 3.**
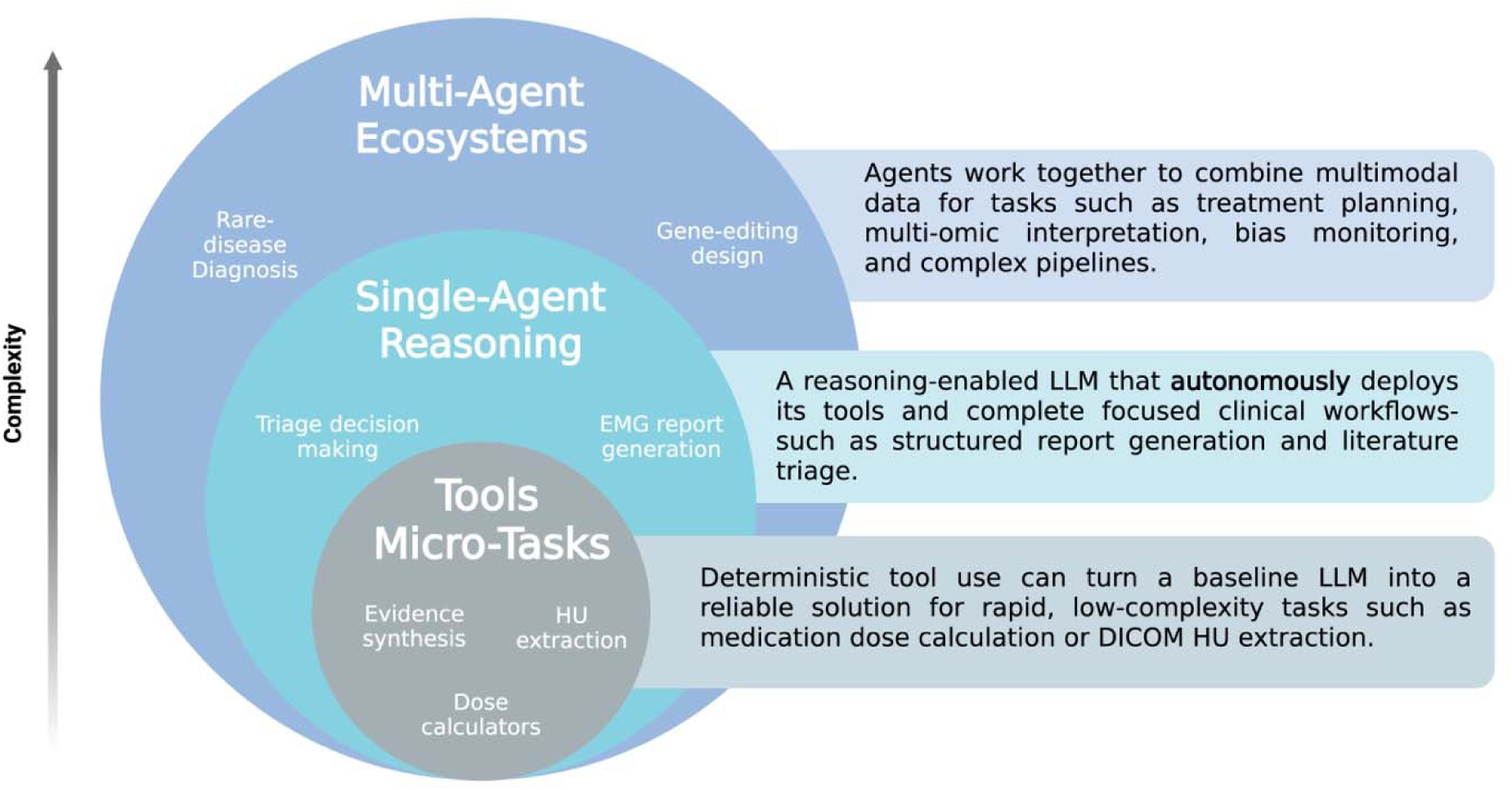
Hierarchical taxonomy of agentic AI architectures in clinical medicine Concentric circles delineate the progressive complexity of agentic systems. *Inner circle*: Tool-based micro-tasks**-** deterministic tool invocation by a baseline LLM for rapid, low-complexity operations (for example, evidence synthesis, dose calculators, DICOM HU extraction). *Middle circle*: Single-agent reasoning**-** an autonomous, reasoning-enabled LLM that self-selects tools to complete an end-to-end clinical workflow (for example, EMG report generation, literature triage). *Outer circle*: Multi-agent ecosystems**-** coordinated networks of specialised, tool-enabled agents that fuse multimodal data to tackle high-stakes, cross-disciplinary problems (for example, rare-disease diagnosis, gene-editing design, post-deployment surveillance). The schematic underpins the three-tier implementation framework proposed in this review.

**Table 3.** AI Agent Architecture Selection Framework for Clinical Medicine.

| Architecture Type | When to Use (Decision Criteria) | Clinical Applications | Implementation Requirements | Key Considerations & Limitations |
| --- | --- | --- | --- | --- |
| **Simple Tool Augmentation**<br>*Direct tool invocation without iteration* | • Single, deterministic operation<br>• Well-defined input/output<br>• No reasoning required<br>• Immediate response needed<br>• Clear success metrics | Medication Dosing<br>Lab Value Conversion<br>DICOM HU Extraction<br>ICD-10 Lookup<br>Drug Interaction Check<br>Risk Score Calculation | **Technical:**<br>• API integration<br>• Minimal compute<br>• No model fine-tuning<br>**Clinical:**<br>• EHR integration<br>• Validated algorithms<br>• Regulatory clearance for calculations | Limited to discrete tasks<br>No complex reasoning<br>Cannot handle ambiguity<br>**Best for:** High-volume, repetitive tasks requiring consistent accuracy<br>**Avoid for:** Tasks requiring clinical judgment or context |
| **Single-Agent Systems**<br>*Autonomous reasoning with tool orchestration* | • Multi-step workflows<br>• Tool selection decisions<br>• Iterative refinement needed<br>• End-to-end task completion<br>• Context preservation required | Radiology Reports<br>EMG Interpretation<br>Literature Triage<br>Clinical Summaries<br>Treatment Planning<br>Discharge Notes | **Technical:**<br>• ReAct/CoT prompting<br>• Memory management<br>• Tool API suite<br>• GPT-4+ class models<br>**Clinical:**<br>• Workflow integration<br>• Audit trails<br>• Human oversight protocols | Context window limits<br>Single perspective<br>Tool timing critical<br>**Best for:** Structured workflows with clear objectives<br>**Optimal tools:** 3-5 specialized tools<br>**Key success factor:** Well-defined task boundaries |
| **Multi-Agent Ecosystems**<br>*Collaborative specialized agents* | • Cross-specialty expertise<br>• Conflicting evidence synthesis<br>• Bias mitigation critical<br>• Complex consensus needed<br>• Interdisciplinary collaboration | Rare Disease Dx<br>Tumor Board<br>Gene Editing<br>Virtual Lab<br>Multi-Omics<br>ICU Management | **Technical:**<br>• Agent orchestration<br>• Consensus mechanisms<br>• Distributed compute<br>• Optimal: 4-5 agents | Coordination overhead<br>Conflicting outputs<br>Error propagation<br>**Best for:** Genuinely interdisciplinary challenges<br>**Caution:** Diminishing returns >5 agents |
|  |  |  | **Clinical:**<br>• Multi-disciplinary protocols<br>• Role definitions<br>• Conflict resolution<br>• Clinical Biases |  |

#### Limitations

This systematic review has several limitations. First, the substantial heterogeneity in clinical domains, outcome metrics, and baseline comparators precluded meaningful quantitative meta-analysis, limiting our ability to provide precise effect estimates.

Second, we restricted peer-reviewed publications, though this decision enhanced quality assurance. Third, the analysis of scaling effects (agent and tool number) is confounded by task heterogeneity and varying comparison baselines. Further, no study provides a comparison of multi-agent to single-agent. Finally, the absence of standardized evaluation frameworks for AI agent systems means performance metrics vary widely across studies, complicating direct comparisons.

#### Future Directions-Emerging Evidence from non-peer review Literature

When looking at the preprint analysis we identified further exploration beyond AI agents’ performance. Microsoft AI study who showed the cost-effectiveness of reducing unnecessary medical tests through a multi-agent framework (MAI-DxO)^24^. Additionally the integration of AI agents into hospitals and EHR.^56,57,77^ With Tsinghua University taking the simulated AI-hospital transforming it into real-time evaluations of a hospital operating entirely on AI agents.^78^ Such advancements could significantly ease the documentation burden on physicians^79,80^ and reducing costs and time.^81^ Future studies should explore prospective validations, create standardized clinical evaluation framework, explore secondary outcome, consensus, task task-architecture matching and ethics in AI agent^82^

## Conclusions

AI agents improve clinical task performance compared with base LLMs, particularly when architecture is matched to task complexity. Tool-augmented and single-agent systems are ready for near-term deployment, while multi-agent frameworks should be limited to high-complexity problems under research conditions. We define AI agents as systems combining iterative reasoning with tool use and/or multi-agent collaboration to guide consistent study and reporting. Future work should emphasize prospective, multi-center trials with real-world data, standardized evaluation, and integration with electronic health records within clear regulatory frameworks.

## Data Availability

The datasets generated during and/or analyzed during the current study are available from the corresponding author on reasonable request.

## Notes

### Competing Interest Statement

The authors have declared no competing interest.

