## Appendix for "AI Agents in Clinical Medicine: A Systematic Review"

#### Table of Contents

***Supplementary Appendix*** **1**

**AI Agents in Clinical Medicine: A Systematic Review** **1**

**Supplementary Methods** **3**
- **S1. Detailed Search Strategy** 3
- **S2. Study Selection Process** 5
- **S3. Data Extraction Protocol** 6
- **S4. Risk of Bias Assessment** 7
- **S5. Statistical Analysis Details** 8

**Supplementary Results** **9**
- **S6. Extended Study Characteristics** 9
- **S7. Non Peer Review Literature Analysis** 10
- **S8. Performance Metrics Analysis** 11

**Supplementary Tables** **12**
- **Supplementary Table 1. QUADAS-AI Risk of Bias Assessment** 12
- **Supplementary Table 2. Preprint Studies Characteristics** 14
- **Supplementary Table 3. Architecture Selection Decision Matrix** 15
- **Supplementary Table 4. Implementation Readiness Assessment** 15
- **Supplementary Table 5. Consensus Mechanisms in Multi-Agent Studies** 16
- **Supplementary Table 6. LLM Backbone Distribution and Performance** 16
- **Supplementary Table 7. Sample Size Distribution by Clinical Task** 16
- **Supplementary Table 8. Frameworks and Implementation Details** 17
- **Supplementary Table 9. Key Performance Improvements by Study** 17
- **Supplementary Table 10. Key Performance Improvements from baseline LLM by Architecture Type** 18

**Supplementary Figures** **19**
- **Supplementary Figure 1. PRISMA Flow Diagram** 19
- **Supplementary Figure 2. System Complexity versus Performance** 20
- **Supplementary Figure 3. Forest Plot of Performance Improvements by Study** 22
- **Supplementary Figure 4. Temporal Evolution of Agentic Architectures** 23
- **Supplementary Figure 5. Landscape of clinical tasks addressed by AI agents** 24
- **Supplementary Figure 6. Agentic AI in clinical medicine-core criteria and exclusions** 26

**Additional Supplementary Information** **28**
- **S9. Code and Data Availability** 28

Mount Sinai

Division of Data-Driven and Digital Medicine (D3M), Icahn School of Medicine at Mount Sinai, New York, USA

**S10. Amendments to Protocol** 28
**S11. Author Contributions** 28
**S12. Protocol Registration and Amendments** 28
**S13. Search Updates** 28
**S14. Data Availability Statement** 29

### Supplementary Methods

#### S1. Detailed Search Strategy

##### S1.1 Database Selection and Rationale

We conducted a comprehensive literature search across three major bibliographic databases:

- **PubMed/MEDLINE**: Primary biomedical literature database
- **Web of Science Core Collection**: Multidisciplinary coverage with citation analytics
- **Scopus**: Broad scientific coverage

Search period: October 1, 2022, to August 5, 2025

##### S1.2 Search Terms and Boolean Operators

**PubMed Search Strategy (N = 4,706 results)**

```
((("large language model"[tiab] OR "language model"[tiab] OR LLM[tiab] OR
ChatGPT[tiab] OR GPT[tiab] OR GPT-3[tiab] OR GPT-4[tiab] OR
"generative AI"[tiab] OR "generative pre-trained transformer"[tiab] OR
"foundation model"[tiab] OR "AI assistant"[tiab] OR chatbot[tiab] OR
Bard[tiab] OR Gemini[tiab] OR Claude[tiab] OR Llama[tiab])
OR
("Artificial Intelligence"[Mesh] OR "Natural Language Processing"[Mesh] OR
"Machine Learning"[Mesh] OR "Computational Linguistics"[Mesh]))

AND

(agentic[tiab] OR agent[tiab] OR "AI agent"[tiab] OR "autonomous agent"[tiab]
OR
autonomous[tiab] OR "tool use"[tiab] OR "tool calling"[tiab] OR
"multi-agent"[tiab] OR LangChain[tiab] OR AutoGen[tiab] OR ReAct[tiab] OR
"assistive"[tiab] OR "prompt engineering"[tiab] OR "Software Agents"[Mesh])

AND

(("Medicine"[Mesh] OR "Clinical Medicine"[Mesh] OR "Health Care"[Mesh] OR
"Diagnosis"[Mesh] OR "Therapeutics"[Mesh] OR
  "Surgery"[Mesh] OR "Genetics"[Mesh] OR "Internal Medicine"[Mesh] OR
"Neurology"[Mesh] OR "Radiology"[Mesh] OR
  "Diagnostic Imaging"[Mesh] OR "Nuclear Medicine"[Mesh] OR
"Cardiology"[Mesh] OR "Nephrology"[Mesh] OR
  "Gastroenterology"[Mesh] OR "Endocrinology"[Mesh] OR "Hematology"[Mesh] OR
"Medical Oncology"[Mesh] OR
```

```
  "Pulmonary Disease"[Mesh] OR "Rheumatology"[Mesh] OR "Infectious
Disease"[Mesh] OR "Immunology"[Mesh] OR
  "Allergy and Immunology"[Mesh] OR "Geriatrics"[Mesh] OR "Dermatology"[Mesh]
OR "Ophthalmology"[Mesh] OR
  "Otorhinolaryngologic Diseases"[Mesh] OR "Urology"[Mesh] OR
"Gynecology"[Mesh] OR "Obstetrics"[Mesh] OR
  "Pediatrics"[Mesh] OR "Psychiatry"[Mesh] OR "Family Practice"[Mesh] OR
"Orthopedics"[Mesh] OR "Dentistry"[Mesh] OR
  "Pathology"[Mesh] OR "Pharmacology"[Mesh] OR "Anesthesiology"[Mesh] OR
"Emergency Medicine"[Mesh] OR
  "Critical Care"[Mesh] OR "Rehabilitation"[Mesh])
 OR medicine[tiab] OR medical[tiab] OR clinical[tiab] OR healthcare[tiab] OR
health[tiab] OR diagnosis[tiab] OR
   treatment[tiab] OR patient[tiab] OR hospital[tiab] OR surgery[tiab] OR
surgical[tiab] OR radiology[tiab] OR imaging[tiab] OR
   cardiology[tiab] OR nephrology[tiab] OR gastroenterology[tiab] OR
endocrinology[tiab] OR hematology[tiab] OR oncology[tiab] OR
   pulmonology[tiab] OR rheumatology[tiab] OR "infectious disease"[tiab] OR
immunology[tiab] OR geriatrics[tiab] OR
   dermatology[tiab] OR ophthalmology[tiab] OR otolaryngology[tiab] OR
urology[tiab] OR gynecology[tiab] OR obstetrics[tiab] OR
   pediatrics[tiab] OR psychiatry[tiab] OR neurology[tiab] OR
orthopedics[tiab] OR dentistry[tiab] OR pharmacology[tiab] OR
   anesthesiology[tiab] OR "emergency medicine"[tiab] OR "critical
care"[tiab] OR rehabilitation[tiab]))


AND

2022/10/01:2025/08/06[dp]
```

**Web of Science Search Strategy (N = 5,844 results)**

```
TS=(("large language model" OR "language model" OR LLM OR ChatGPT OR GPT OR
GPT-3 OR GPT-4 OR "generative AI" OR "generative pre-trained transformer" OR
"foundation model" OR "AI assistant" OR chatbot OR Bard OR Gemini OR Claude
OR Llama)
 AND

 (agentic OR agent OR "AI agent" OR "autonomous agent" OR autonomous OR "tool
use" OR "tool calling" OR "multi-agent" OR LangChain OR AutoGen OR ReAct OR
assistive OR "prompt engineering")
 AND

 (medicine OR medical OR clinical OR healthcare OR health OR diagnosis OR
treatment OR patient OR hospital OR surgery OR surgical OR radiology OR
imaging OR cardiology OR nephrology OR gastroenterology OR endocrinology OR
hematology OR oncology OR pulmonology OR rheumatology OR "infectious disease"
OR immunology OR geriatrics OR dermatology OR ophthalmology OR otolaryngology
OR urology OR gynecology OR obstetrics OR pediatrics OR psychiatry OR
neurology OR orthopedics OR dentistry OR pharmacology OR anesthesiology OR
"emergency medicine" OR "critical care" OR rehabilitation) )
AND
```

```
 PY=(2022-2025)
```

**Scopus Search Strategy (N = 261 results)**

```
(("large language model" OR LLM OR ChatGPT OR GPT-3 OR GPT-4 OR
Claude OR Deepseek OR Gemini)
AND
("tool use" OR "tool calling" OR "AI Agent" OR "multi-agent" OR
"autonomous agent" OR LangChain OR AutoGen OR ReAct)
AND
(medicine OR medical OR clinical OR healthcare OR diagnosis OR
treatment OR patient OR hospital))
```

**Total initial results across all databases: 10,811 After deduplication: 4,927 unique records**

#### S2. Study Selection Process

##### S2.1 Screening Protocol

###### Phase 1: Title and Abstract Screening

- Two independent reviewers (A.G., M.O.) screened all 6023 records
- Covidence systematic review software utilized for management
- 5769 records excluded at title/abstract stage
- 254 records advanced to full-text review
- Inter-rater reliability: Cohen's κ = 0.93
- Inclusion criteria applied:
    1. AI agent system described
    2. Human health-related application
    3. Quantitative performance metrics reported
    4. Peer-reviewed publication in English

###### Phase 2: Full-Text Assessment

- 254 articles underwent full-text review
- 234 articles excluded (see exclusion tracking below)
- 20 articles included in final analysis
- Disagreements resolved through discussion (n=9)
- Third reviewer (E.K.) arbitration required (n=3 cases)

##### S2.2 Exclusion Tracking

| Exclusion Reason | Number of Studies | Examples |
|---|---|---|

| | | |
|---|---|---|
| **Non-agentic single-pass LLM implementations** | 181 | Studies using LLM without tools and iteration |
| **Simulation or benchmark-only studies** | 31 | No clinical validation or real-world tasks |
| **Studies lacking comparative performance metrics** | 22 | Descriptive studies without performance data |
| **Total excluded at full-text stage** | **234** | |

#### S3. Data Extraction Protocol

##### S3.1 Reviewer Training and Calibration

1. **Initial Training Phase**
   - Both reviewers (A.G., M.O.) are well trained in the field and had standardized training module
   - Training materials included:
     - Definitions of AI agents vs. non-agentic systems (based on ReACT prompting, tool-calling, multi-agent collaboration)
     - Clinical domain categorization guidelines (9 categories identified)
     - Performance metric extraction standards (accuracy, F1 score, ROUGE-L, custom metrics)
2. **Pilot Extraction**
   - 5 randomly selected studies for calibration
   - Independent extraction followed by consensus discussion
   - Form refinement based on pilot findings
   - Agreement rate post-calibration: >90%
3. **Quality Assurance**
   - 20% of extractions (4 studies) cross-validated
   - Daily consensus meetings during extraction period (June-July 2025)
   - Discrepancy log maintained
   - Third reviewer (E.K.) arbitration required for 3 studies

##### S3.2 Data Extraction Form

##### Section A: Study Characteristics

- First author, year, country
- Funding source
- Conflict of interest declarations
- Study registration (if applicable)

##### Section B: Technical Specifications

- Base LLM(s) and version
- Agentic framework (commercial/custom)
- Agent architecture classification:
    - Single agent with tools
    - Multi-agent without tools
    - Multi-agent with tools
- Number of agents (if multi-agent)
- Tool specifications:
    - Tool types (API, database, computation, etc.)
    - Tool count
    - Tool integration method

**Section C: Clinical Application**

- Clinical domain (primary and secondary)
- Task complexity classification
- Data source characteristics:
    - Real-world vs. synthetic
    - Sample size
    - Data completeness

**Section D: Performance Metrics**

- Primary outcome measure
- Baseline comparator type
- Absolute performance values
- Relative improvement calculations
- Statistical significance (if reported)
- Confidence intervals/standard errors

#### S4. Risk of Bias Assessment

##### S4.1 QUADAS-AI Tool Adaptation

We adapted the QUADAS-AI (Quality Assessment of Diagnostic Accuracy Studies for AI) tool for agentic systems evaluation:

**Domain 1: Patient/Data Selection**

- High risk indicators:
    - Synthetic data only
    - Single center without justification
    - Non-consecutive or convenience sampling
    - Sample size <100 without power calculation

**Domain 2: Index Test (AI System)**

- High risk indicators:
  - Unclear model version or parameters
  - No description of prompt engineering
  - Threshold not prespecified
  - Post-hoc optimization on test set

**Domain 3: Reference Standard**

- High risk indicators:
  - Single annotator without expertise verification
  - Automated labeling without validation
  - Circular reasoning (LLM validating LLM)
  - Unclear ground truth establishment

**Domain 4: Flow and Timing**

- High risk indicators:
  - Different datasets for agent vs. baseline
  - Temporal mismatch in data collection
  - Selective outcome reporting
  - Missing data not addressed

##### S4.2 Reviewer Calibration for Bias Assessment

- Initial agreement rate: 90%
- Post-calibration agreement: 95%
- Arbitration required: 1 studies

#### S5. Statistical Analysis Details

##### S5.1 Handling of Heterogeneous Metrics

Given the diversity of outcome measures, we employed the following standardization approach:

1. **Accuracy-based metrics**: Converted to percentage scale
2. **F1 scores**: Maintained as reported (0-1 scale)
3. **Custom scores**: Normalized to 0-100 scale where possible
4. **Relative improvements**: Calculated as (Agent - Baseline)/Baseline × 100%

##### S5.2 Subgroup Analyses

Stratification variables:

- Architecture type (3 categories)
- Clinical domain (9 categories)
- Data type (real-world vs. synthetic)
- Baseline comparator (LLM vs. human)

---

### Supplementary Results

#### S6. Extended Study Characteristics

##### S6.1 Study Distribution by Year and Design

| Publication Year | Number of Studies | Study Designs |
|---|---|---|
| **2024** | 2 | Comparative (1), Simulation (1) |
| **2025 (through August)** | 18 | Comparative (15), RCT (1), Retrospective (1), Simulation (1) |

##### S6.2 Clinical Domain Distribution

| Clinical Domain | Number of Studies | Percentage | Median Sample Size |
|---|---|---|---|
| **Diagnosis & Prognosis** | 8 | 40% | 238 (IQR: 16-302) |
| **Evidence Synthesis** | 5 | 25% | 272 (IQR: 50-500) |
| **Treatment Planning** | 3 | 15% | 37 (IQR: 17-92) |
| **Clinical Operations** | 2 | 10% | 5,050 (range: 100-10,000) |
| **Genomics Applications** | 2 | 10% | 689 (range: 288-1,106) |
| **Medical Education** | 1 | 5% | 1,062 |

#### S7. Non Peer Review Literature Analysis

##### S7.1 Search Strategy for Preprints

Databases searched:

- arXiv (cs.AI, cs.CL sections)
- medRxiv
- bioRxiv

Search terms: Without search strategy Date range: Same as primary search (October 1, 2022 - August 5, 2025)

##### S7.2 Selection Criteria for Narrative Synthesis

Inclusion criteria for preprint discussion:

1. High technical quality (determined by two reviewers)
2. Novel architectural contributions
3. Innovate solution
4. Clinical applicability

**Selected Preprints for Detailed Analysis (n=5):**

| Study | Repository | Citation Count* | Key Innovation | Sample Size |
|---|---|---|---|---|
| **Li et al. (Agent Hospital)** | arXiv | 170 | Evolvable agents with learning | 339 diseases, 32 departments |
| **Shi et al. (EHRAgent)** | arXiv | 83 | Code-empowered tabular reasoning | MIMIC-III, eICU datasets |
| **Zuo et al. (KG4Diagnosis)** | arXiv | 15 | Knowledge graph + multi-agent | 362 common diseases |
| **Nori et al. (MAI-DxO)** | arXiv | 4 | Cost-optimized diagnosis | 304 NEJM cases |
| **Chen et al. (RadFabric)** | arXiv | 0 | Modular radiology agents | Multiple CXR benchmarks |

*Citation counts as of August 2025

#### S8. Performance Metrics Analysis

##### S8.1 Distribution of Performance Improvements

| Architecture Type | Median Absolute Improvement | Median Relative Improvement | IQR (Absolute) | Range (Absolute) | N Studies |
|---|---|---|---|---|---|
| **Single Agent + Tools** | 48.0 pp | 165.6% | 36.0-56.9 pp | 29.6-61.7 pp | 8 |
| **Multi-Agent Only** | 14.3 pp | 58.3% | 4.7-72.6 pp | 4.07-28.0 pp | 5 |
| **Multi-Agent + Tools** | 7.28 pp* | 29.7% | -0.11-49 pp | -0.24-49 pp | 7 |
| **Overall** | **28.0 pp** | **61.0%** | **-** | **-0.24-61.7 pp** | **20** |

*Note: Includes different metric types and different baseline comparison (base-LLM and physicians). Two studies (Yang 2025, Xu et al.) had no baseline comparison and were excluded from calculations.

**Key Findings After Data Correction:**

- Single-agent + tools architectures show the highest median relative improvement (165.6%)
- Multi-agent systems show more modest improvements when tools are added
- Two studies showed agent performance worse than comparator (Gorenshtein 2025a: -0.11% vs Sonar-pro; Gorenshtein 2025c: -25.5% vs physicians)

##### S8.2 Tool Utilization Analysis

| Tool Type | Frequency | Studies Using | Median Performance Gain |
|---|---|---|---|
| **Web Search/APIs** | 8 (40%) | Gorenshtein 2025a, Ferber, Qu, Pickard, Xu, Yang 2024, Low, Wang 2025a | 42.3% |
| **RAG/Retrieval** | 7 (35%) | Goodell, Gorenshtein 2025b/c, Ferber, Woo, Mejia, Yang 2024 | 36.5% |
| **Code Execution** | 5 (25%) | Goodell, Ferber, Pickard, Yang 2025, Low | 53.5% |

| | | | |
|---|---|---|---|
| **Vision/Image Analysis** | 3 (15%) | Ferber, Chen (preprint), RadFabric (preprint) | 56.9% |
| **Database/EHR Integration** | 6 (30%) | Gorenshtein 2025c, Pickard, Wang 2025a, Xu, Mejia, Shi (preprint) | 29.6% |
| **Domain-specific Tools** | 4 (20%) | Qu (Primer3), Swanson (ESMFold), Wang 2025b (optimization), Mejia (ML models) | 31.2% |

---

### Supplementary Tables

#### Supplementary Table 1. QUADAS-AI Risk of Bias Assessment

| Study | Patient Selection | Index Test | Reference Standard | Flow & Timing | Overall Risk | Key Concern |
|---|---|---|---|---|---|---|
| **Chen et al.** | ⊕ Low | ⊕ Low | ⊕ Low | ⊕ Low | ⊕ Low | None |
| **Goodell et al.** | ⊖ High | ⊕ Low | ⊕ Low | ⊕ Low | ⊖ High | Synthetic data |
| **Omar et al.** | ⊖ High | ⊕ Low | ⊕ Low | ⊕ Low | ⊖ High | Exam questions only |
| **Gorenshtein 2025a** | ⊖ High | ⊕ Low | ⊖ High | ⊕ Low | ⊖ High | Fictional queries |
| **Gorenshtein 2025b** | ⊕ Low | ⊕ Low | ⊕ Low | ⊕ Low | ⊕ Low | Single center retrospective |
| **Gorenshtein 2025c** | ⊕ Low | ⊕ Low | ⊖ High | ⊕ Low | ⊖ High | Custom score |
| **Ferber et al.** | ⊖ High | ⊕ Low | ⊖ High | ⊖ High | ⊖ High | Small synthetic cohort |
| **Qu et al.** | ⊕ Low | ⊕ Low | ⊕ Low | ⊕ Low | ⊕ Low | Wet-lab validation |
| **Pickard et al.** | ⊖ High | ⊕ Low | ⊕ Low | ⊕ Low | ⊖ High | Limited datasets |
| **Woo et al.** | ⊖ High | ⊕ Low | ⊕ Low | ⊕ Low | ⊖ High | Guidelines only |
| **Swanson et al.** | ⊕ Low | ⊕ Low | ⊕ Low | ⊕ Low | ⊕ Low | Lab validation |

| | | | | | | |
|---|---|---|---|---|---|---|
| **Mejia et al.** | ⊕ Low | ⊕ Low | ⊖ High | ⊕ Low | ⊕ Low | Limited validation |
| **Wang 2025a** | ⊕ Low | ⊕ Low | ⊕ Low | ⊕ Low | ⊕ Low | Curated datasets |
| **Yang 2025** | ⊖ High | ? Unclear | ? Unclear | ⊕ Low | ⊖ High | Limited validation |
| **Altermatt et al.** | ⊖ High | ⊕ Low | ⊕ Low | ⊕ Low | ⊖ High | Exam questions |
| **Xu et al.** | ⊖ High | ⊕ Low | ⊕ Low | ⊕ Low | ⊖ High | Article analysis only |
| **Wang 2025b** | ⊖ High | ⊕ Low | ⊕ Low | ⊕ Low | ⊖ High | Small cohort |
| **Ke et al.** | ⊖ High | ⊕ Low | ⊕ Low | ⊕ Low | ⊖ High | Case reports only |
| **Yang 2024** | ⊖ High | ⊕ Low | ⊕ Low | ⊕ Low | ⊖ High | Case reports |
| **Low et al.** | ⊖ High | ? Unclear | ⊕ Low | ⊕ Low | ⊖ High | Limited details |

#### Supplementary Table 2. Preprint Studies Characteristics

| Study (First author, Year) | Title | Preprint date | Domain / Task | Agentic setup | Data / Evaluation | Key outcomes | Notes / Limitations |
|---|---|---|---|---|---|---|---|
| Li et al., 2025 | Agent Hospital: A Simulacrum of Hospital with Evolvable Medical Agents | 2025-01-17 | End-to-end hospital simulacrum; exam selection, diagnosis, treatment planning (virtual + MedQA) | Multi-agent (patients, nurses, doctors); evolvable agents; SEAL paradigm | Virtual world across 32 departments / 339 diseases; real-world: MedQA (GPT-4o base) | Doctor agents improve with experience (e.g., cardiology case 9%→82%); outperformed MedAgents/COT/MedPrompt on MedQA | Relies on simulated training data; proposes alignment to real-world; ethical considerations discussed |
| Shi et al., 2024 | EHRAgent: Code Empowers LLMs for Few-shot Complex Tabular Reasoning on EHRs | 2024-10-04 | EHR multi-tabular QA and analytics | Single agent with tools (code-generation + execution); long-term memory; interactive debugging | MIMIC-III, eICU, TREQS; GPT-4 (0613) base; success rate (SR), completion rate (CR) | +19.92% (MIMIC-III), +12.41% (eICU), +29.60% (TREQS) SR over strongest baselines; ablations highlight interactive coding | More LLM calls (cost) vs open-loop; privacy/ethics considerations; plans to add open LLMs |
| Zuo et al., 2025 | KG4Diagnosis: A Hierarchical Multi-Agent LLM Framework with Knowledge Graph Enhancement for Medical Diagnosis | 2025-01-03 | Diagnostic reasoning with knowledge graph | Hierarchical multi-agent (GP triage + specialist agents) with automated KG construction | Scope: 362 common diseases; modular framework; benchmarks noted in paper | Framework paper; emphasizes KG constraints + multi-agent verification to mitigate hallucinations; implementation guidelines provided | Primarily architectural; limited quantitative metrics in abstract |
| Nori et al., 2025 | Sequential Diagnosis with Language Models | 2025-06-30 | Interactive diagnostic agents; sequential reasoning benchmark | MAI-DxO orchestrator (panel-of-physicians simulation) atop various LMs | SDBench (304 NEJM CPC cases); measures accuracy + estimated cost | With o3: 80% accuracy (4× physicians) and −20% cost; up to 85.5% accuracy in max-accuracy mode; gains generalize across model families | Benchmark-driven; clinical deployment not studied |
| Chen et al., 2025 | RadFabric: Agentic AI System with Reasoning Capability for Radiology | 2025-06-17 | Radiology (CXR) — detection, localization, reasoning, reporting | Multi-agent, multimodal (specialized CXR vision agents + anatomical interpretation + LLM reasoning) | Multiple CXR tasks; overall and per-pathology accuracy reported | Overall diagnostic accuracy 0.799; near-perfect fracture detection (1.000 accuracy); outperforms traditional systems (0.229–0.527) | Focus on CXR; broader generalization to other modalities not evaluated here |

#### Supplementary Table 3. Architecture Selection Decision Matrix

| Task Characteristic | Simple Tools | Single Agent | Multi-Agent | Key Decision Factor |
|---|---|---|---|---|
| Requires iterative reasoning? | **No** | **Yes** | **Yes** | If no → Simple Tools sufficient |
| Multiple tool decisions? | **No** | **Yes** | **Yes** | Dynamic tool selection → Agent needed |
| Cross-specialty expertise? | **No** | **No** | **Yes** | Interdisciplinary → Multi-agent |
| Conflicting evidence? | **No** | **Limited** | **Yes** | Consensus needed → Multi-agent |
| Real-time response (<1s)? | **Fast-But depend on tool** | **Moderate** | **Slow** | Speed critical → Simple tools |
| Cost sensitivity?* | **Low cost** | **Moderate** | **High cost** | Budget constraints favor simpler architectures |

*per query and tokens (reasoning model consume reasoning tokens)

#### Supplementary Table 4. Implementation Readiness Assessment

| Readiness Factor | Simple Tools | Single Agent | Multi-Agent |
|---|---|---|---|
| Technical Infrastructure | Basic API access | Cloud compute, memory management | Distributed systems, orchestration |
| Data Requirements | Structured inputs only | Multi-modal data handling | Complex data fusion & sharing |
| Model Requirements | Any LLM/SLM or specialized tool | High preforming model | Multiple specialized models (High reasoning model and multiple small parameter/SLM models) |
| Clinical Integration | Point solution | Workflow integration | Multi-department coordination |
| Maintenance Complexity | Low - API updates only | Medium - Prompt engineering | High - Agent coordination |

#### Supplementary Table 5. Consensus Mechanisms in Multi-Agent Studies

| Consensus Type | Studies (n=11) | Description | Performance Impact |
|---|---|---|---|
| **Supervisor Coordination** | 4 (36.4%) | Chen et al., Swanson et al., Wang 2025b, Ke et al. | Central orchestrator agent manages workflow |
| **Sequential Processing** | 5 (45.5%) | Gorenshtein 2025a/b/c, Qu et al., Omar et al. | Agents process in defined order with hand-offs |
| **Majority Voting** | 1 (9.1%) | Altermatt et al. | Democratic voting on outputs |
| **Custom/Hybrid** | 1 (9.1%) | Mejia et al. | Resource matching algorithm |

#### Supplementary Table 6. LLM Backbone Distribution and Performance

| LLM Family | Studies Using | As Single Agent | As Multi-Agent | Median Uplift |
|---|---|---|---|---|
| **GPT-4 Family** | 15 (75%) | 8 | 7 | +43.7% |
| **- GPT-4** | 7 | 4 | 3 | +42.3% |
| **- GPT-4o** | 6 | 3 | 3 | +41.0% |
| **- GPT-4-Turbo** | 2 | 1 | 1 | +28.0% |
| **Llama-3 Family** | 4 (20%) | 2 | 2 | +42.5% |
| **Claude-3 Opus** | 4 (20%) | 2 | 2 | +11.5% |
| **Gemini-1.5 Pro** | 4 (20%) | 1 | 3 | +26.4% |
| **Other Models** | 3 (15%) | 1 | 2 | +53.0% |

*Note: Some studies tested multiple models

---

#### Supplementary Table 7. Sample Size Distribution by Clinical Task

| Clinical Task Type | Studies | Median Sample Size | Range | Data Type |
|---|---|---|---|---|
| **Clinical Cases** | 5 | 238 | 16-302 | Mixed (real and synthetic) |
| **Multiple Choice Questions** | 2 | 2,560 | 1,062-4,058 | Exam questions |
| **Clinical Reports/EMG** | 2 | 219 | 200-219 | Real patient data |

| | | | | |
|---|---|---|---|---|
| **Evidence Synthesis Queries** | 4 | 275 | 50-500 | Synthetic queries |
| **Patient Data (actual)** | 2 | 58.5 | 17-100 | Real EHR data |
| **Computational Vignettes** | 1 | 10,000 | - | Synthetic |
| **Genomic/Biological Datasets** | 4 | 690 | 8-1,106 | Mixed |

#### Supplementary Table 8. Frameworks and Implementation Details

| Framework | Studies Using | Open Source | Customization Level |
|---|---|---|---|
| **AutoGen (Microsoft)** | 4 | Yes | Moderate - extensive prompt engineering |
| **LangChain** | 4 | Yes | High - custom chains and tools |
| **Custom Implementation** | 12 | Varies | Complete - built from scratch |
| **CrewAI** | 0 | - | - |
| **Not Specified** | | Unknown | Unknown |

#### Supplementary Table 9. Key Performance Improvements by Study

| Study | Agent Performance | Baseline | Absolute Improvement | Relative Improvement (%) | Comparison Type |
|---|---|---|---|---|---|
| **Goodell et al.** | 95.2% (GPT-4o) | 36.1% | +59.1 pp | +163.7% | vs. Base LLM |
| **Ferber et al.** | 87.2% (GPT-4) | 30.3% | +56.9 pp | +187.8% | vs. Base LLM |
| **Yang 2024** | 0.85 F1 | 0.32 F1 (GPT-3.5) | +0.53 | +165.6% | vs. Base LLM |
| **Qu et al.** | 99% accuracy | 50% (GPT-4o) | +49 pp | +98.0% | vs. Base LLM |
| **Low et al.** | 58% accuracy | 10% (ChatGPT) | +48 pp | +480.0% | vs. ChatGPT |
| **Pickard et al.** | 90.59 ± 18.46 | 28.89 ± 19.44 | +61.7 pp | +213.5% | vs. GPT-4 base |
| **Woo et al.** | 95% accuracy | 59% | +36 pp | +61.0% | vs. Base LLM |
| **Gorenshtein 2025b (INSPIRE)** | 92.2% accuracy | 62.6% | +29.6 pp | +47.3% | vs. Base LLM |
| **Chen et al.** | 34% accuracy | 19.7% | +14.3 pp | +72.6% | vs. Base LLM |

| | | | | | |
|---|---|---|---|---|---|
| **Omar et al. (ICE)** | 74.03% accuracy | 60.2% | +13.83 pp | +23.0% | vs. Base LLM |
| **Wang 2025a** | 0.310 ROUGE-L | 0.239 | +0.071 | +29.7% | vs. Base LLM |
| **Wang 2025b** | 64.75 Gy (D95) | 57.47 Gy | +7.28 Gy | +12.7% | vs. ECHO planner |
| **Gorenshtein 2025a (LITERAS)** | 99.82% accuracy | 99.93% (Sonar-pro) | -0.11 pp | -0.11% | vs. Sonar-pro |
| **Altermatt et al.** | 89.97% accuracy | 85.9% | +4.07 pp | +4.7% | vs. Base LLM |
| **Ke et al.** | 76% accuracy | 48% (physicians) | +28 pp | +58.3% | vs. Physicians |
| **Gorenshtein 2025c** | 0.70 ± 0.26 | 0.94 ± 0.13 (physicians) | -0.24 | -25.5% | vs. Physicians |
| **Mejia et al.** | 0.71 (Qwen-32b) | 0.45 | +0.26 | +57.8% | vs. Base LLM |
| **Swanson et al.** | 2/92 improved | 0/92 baseline | +2 nanobodies | N/A | vs. Wild-type |
| **Yang 2025** | N/A | N/A | N/A | N/A | No comparison |
| **Xu et al.** | 0.825 accuracy | N/A | N/A | N/A | No baseline |

**Note**: Some studies used different metrics or custom scores. For Gorenshtein 2025a (LITERAS), the agent actually performed slightly worse than Sonar-pro but better than base ChatGPT-4o-mini (35.6% for non-academic sources).

#### Supplementary Table 10. Key Performance Improvements from baseline LLM by Architecture Type

| Architecture Type | Number of Studies | Median | Mean | Range | IQR |
|---|---|---|---|---|---|
| **Single Agent + Tools** | 7 | 53.0% | 48.9% | 7.1–61.7% | 36.0–59.1% |
| **Multi-Agent Only** | 4 | 14.05% | 27.05% | 4.1–76.0% | 8.95–45.15% |
| **Multi-Agent + Tools** | 5 | 17.2% | 18.3% | 3.5–49.0% | 4.125–39.3% |
| **Overall** | **16** | **36.0%** | **33.5%** | **3.5–76.0%** | **7.1–56.9%** |

### Supplementary Figures

#### Supplementary Figure 1. PRISMA Flow Diagram

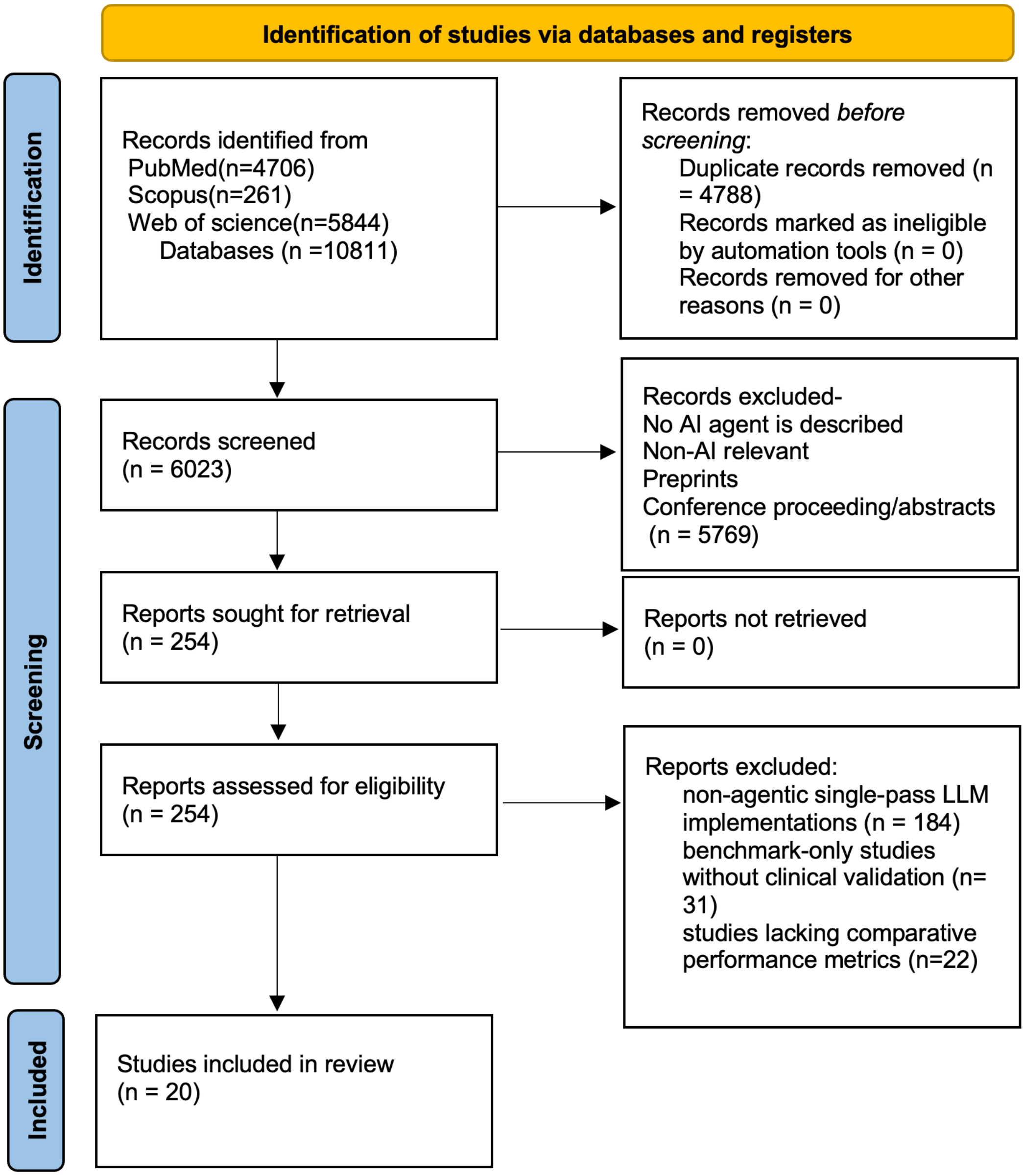

#### Supplementary Figure 2. System Complexity versus Performance

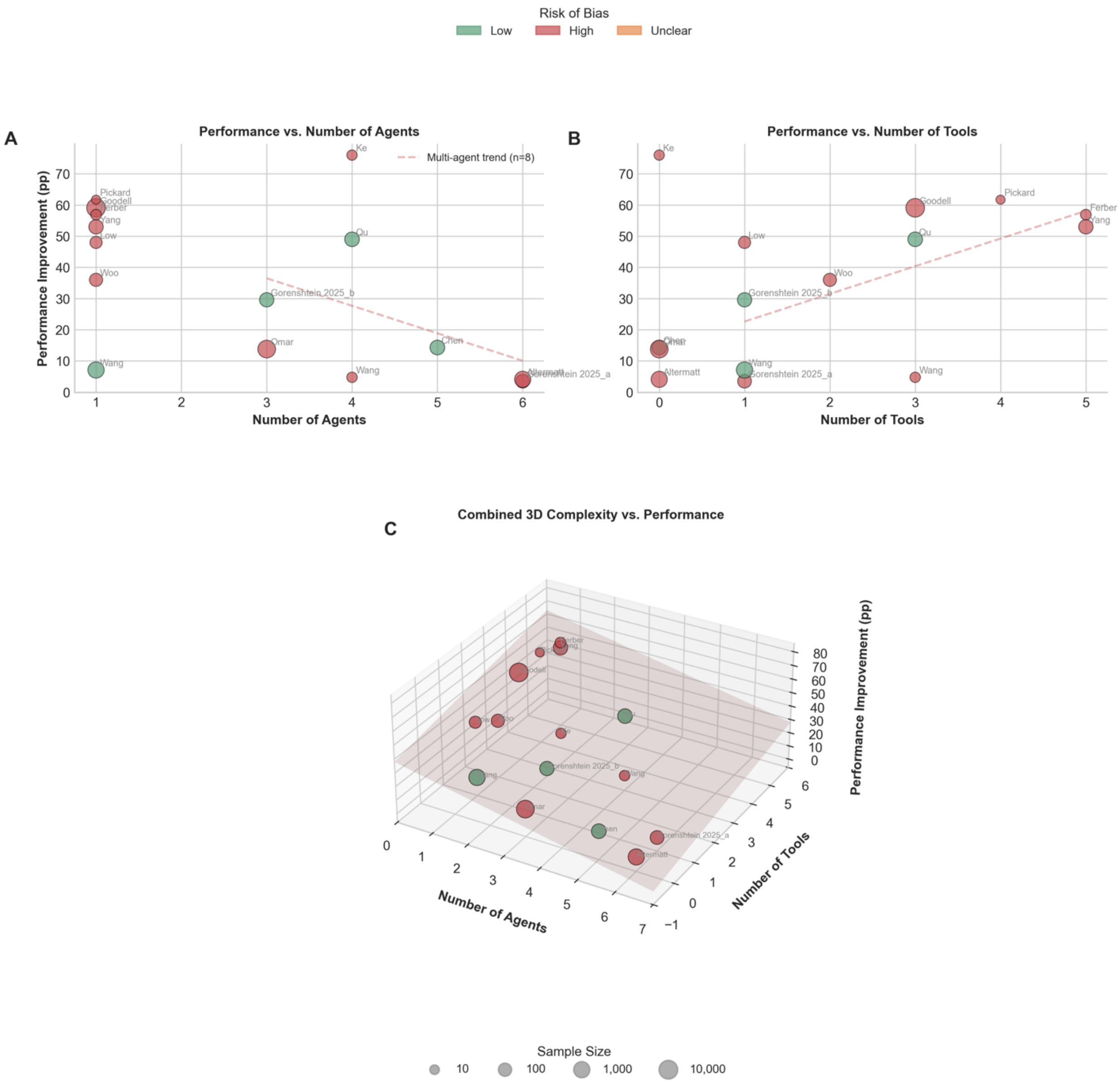


*A,* Scatter plot of performance improvement (pp) of AI Agent LLM version over baseline-LLMs per study; points are sized by sample size and coloured by QUADAS-AI risk of bias (green =

low, red = high, yellow = unclear). Dashed line: linear trend (β = -8.815, $R^2$ = 0.162) showing diminishing returns beyond ~5 agents. *B,* Performance improvement versus number of external tools; trend indicates modest positive association (β = 8.869, $R^2$ = 0.377). *C,* 3-D bubble plot integrating agent count (x), tool count (y) and performance gain (z). Surfaces highlight the inverted-U relationship between architectural complexity and benefit, supporting optimisation rather than maximal scaling of agents and tools ($\beta_{agents}$ = -5.010, $\beta_{tools}$ = 3.551, $R^2$ = 0.328).

### Supplementary Figure 3. Forest Plot of Performance Improvements by Study

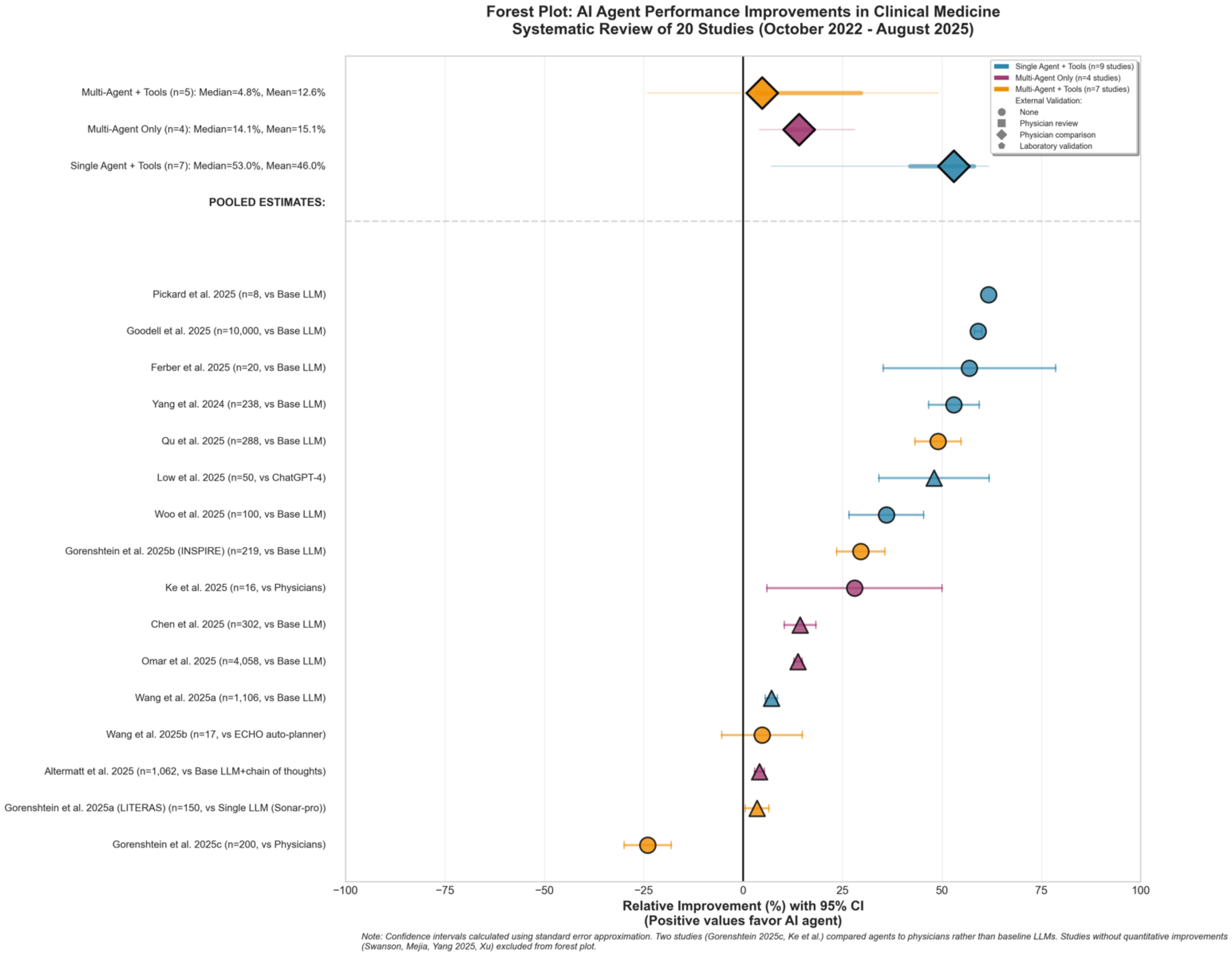

#### Supplementary Figure 4. Temporal Evolution of Agentic Architectures

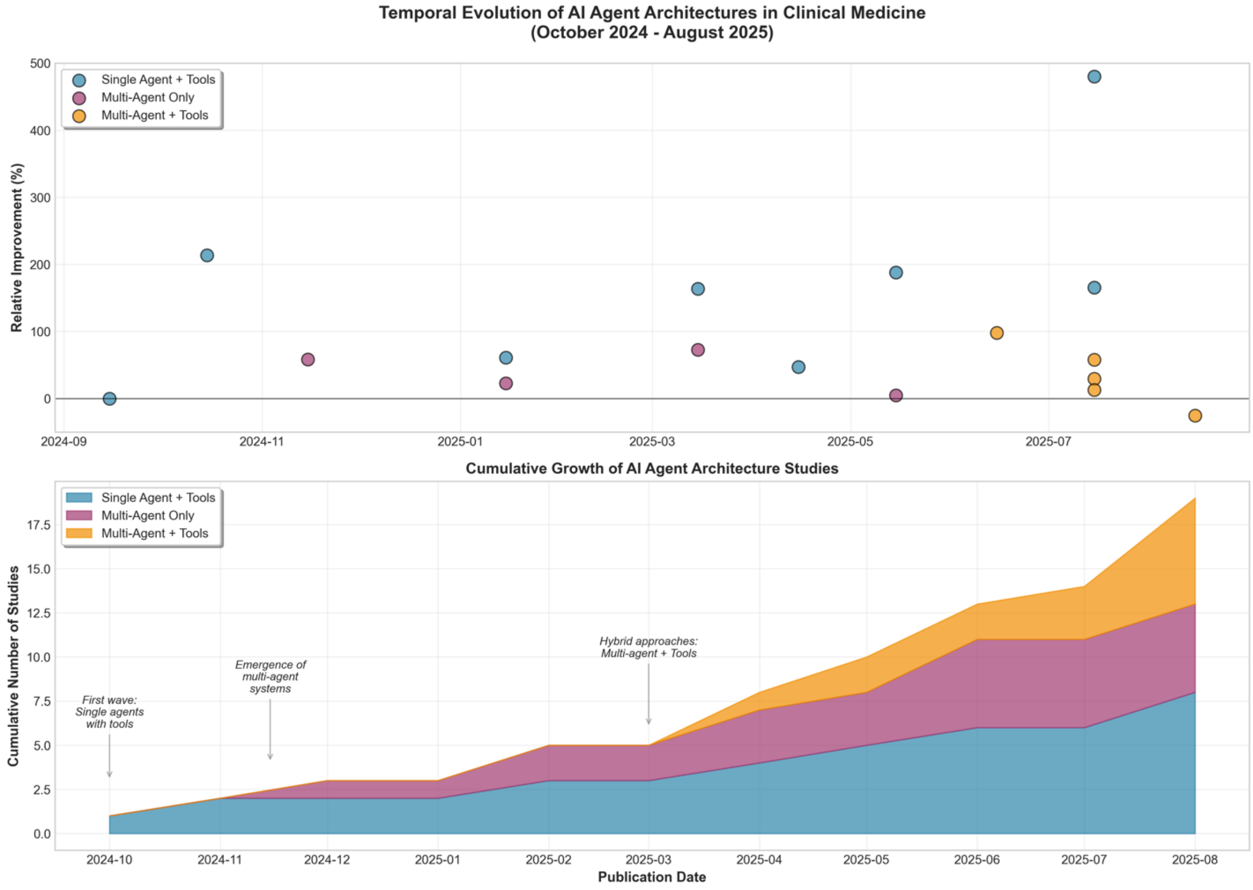


Mount Sinai

Division of Data-Driven and Digital Medicine (D3M), Icahn School of Medicine at Mount Sinai, New York, USA

**Supplementary Figure 5. Landscape of clinical tasks addressed by AI agents**

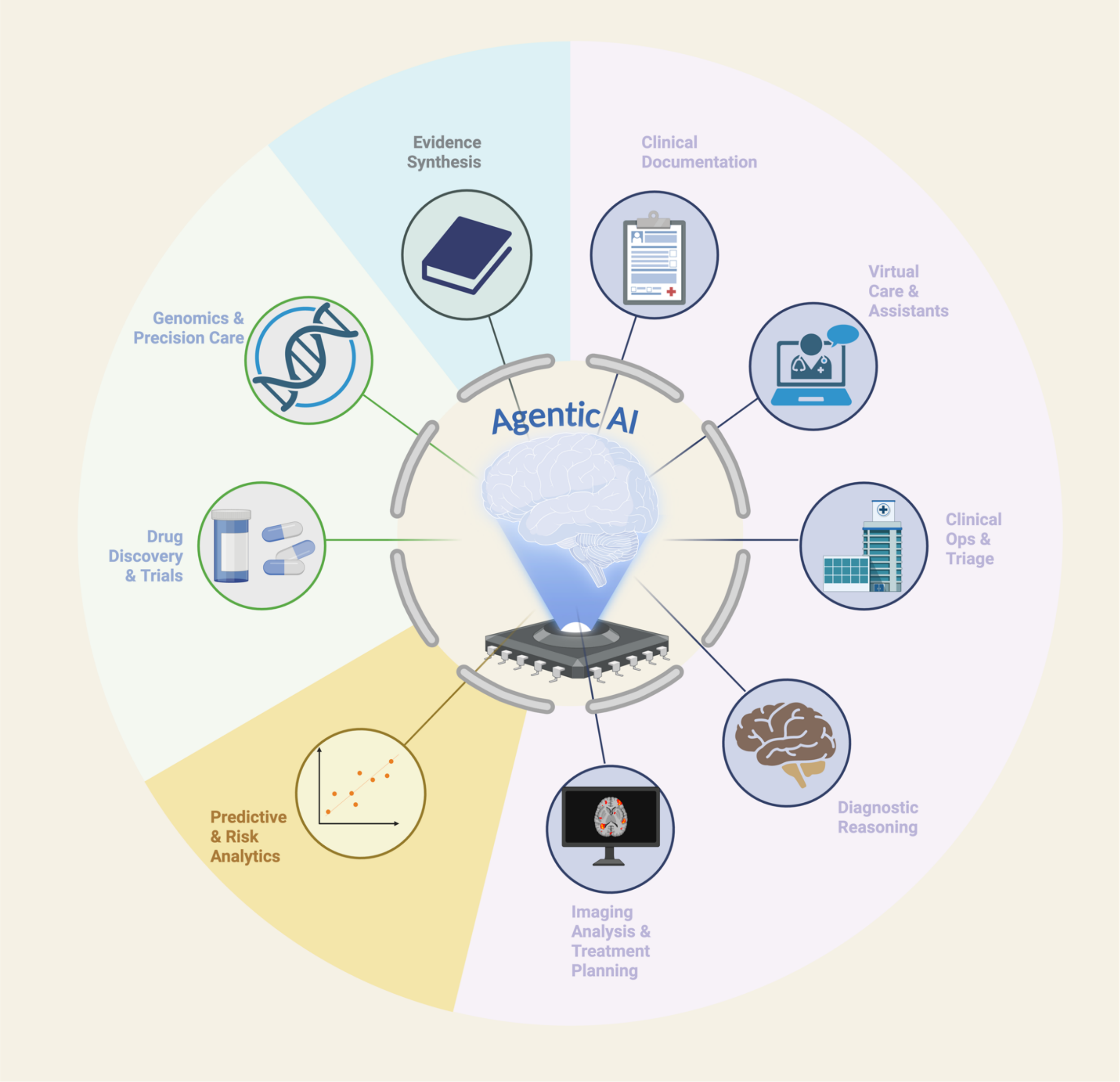


A radial wheel maps the nine principal domains in which agentic systems have been explored to date. Icons illustrate: diagnostics & reasoning, predictive analytics, genomics & precision medicine, clinical operations & triage, evidence synthesis, drug discovery, documentation & administration, virtual care & assistants and imaging analysis/treatment planning. The central robot symbol denotes the AI agent, emphasising that a single architectural paradigm can be adapted across the continuum of patient care.

**Supplementary Figure 6. Agentic AI in clinical medicine-core criteria and exclusions**

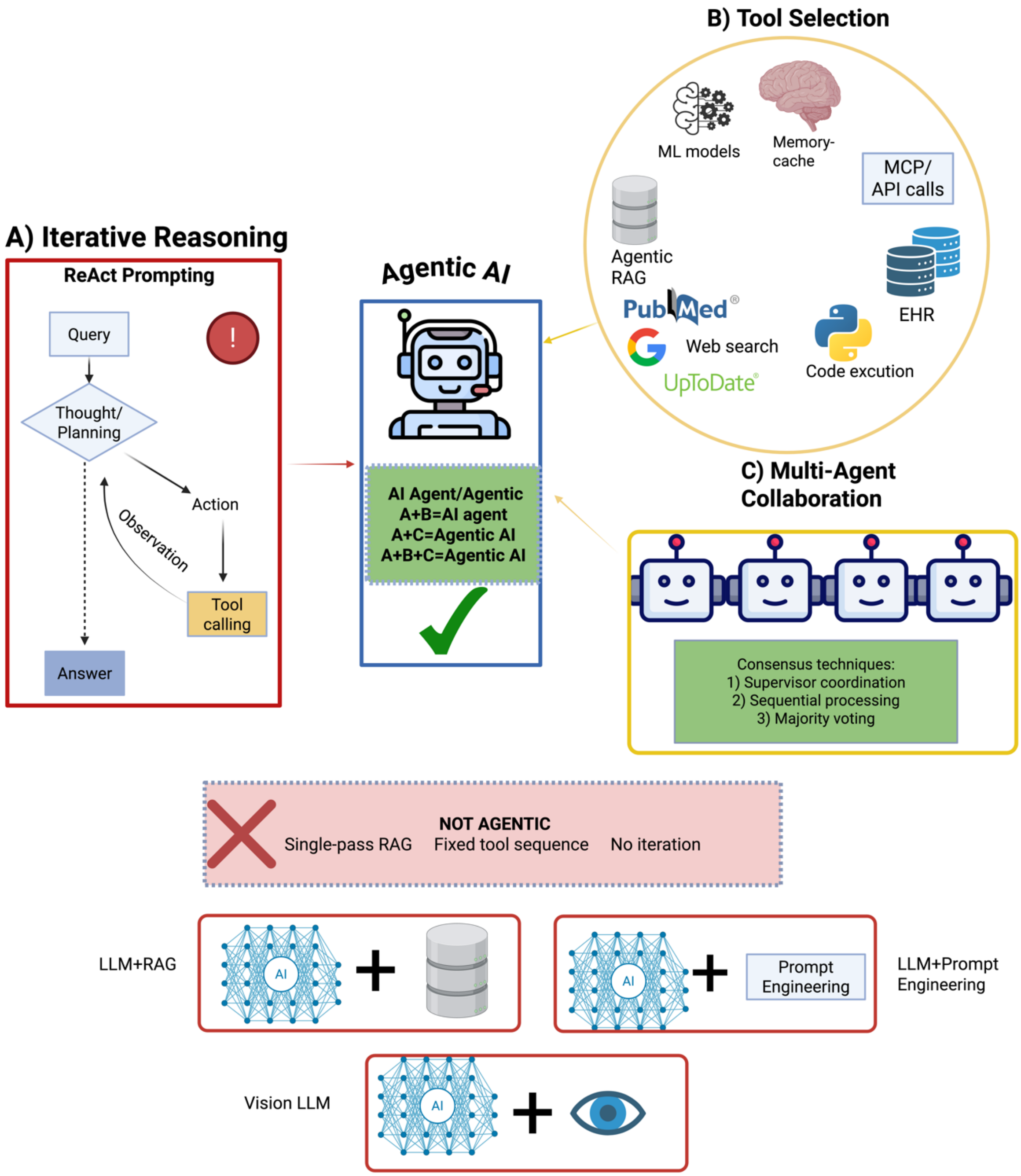


Mount Sinai
Division of Data-Driven and Digital Medicine (D3M), Icahn School of Medicine at Mount Sinai, New York, USA

(A) Iterative reasoning (mandatory): planning–action–observation loops (e.g., ReAct) that update the plan based on feedback. (B) Tool selection: dynamic orchestration of external tools—retrieval/web search, EHR queries, APIs/MCP, memory/cache, code execution, and other ML models—chosen conditionally during the loop. (C) Multi-agent collaboration: coordinated agents using explicit consensus schemes (supervisor/decomposer, sequential hand-offs, or majority voting). Systems implementing (A) plus (B) and/or (C) are classified as agentic. Non-agentic baselines (bottom) include single-pass RAG, fixed tool chains, and prompt-engineering or vision-only models without iteration or dynamic decisions. This operational definition guided study inclusion/exclusion and analytic stratification in the systematic review.

### Additional Supplementary Information

#### S9. Code and Data Availability

- Systematic review protocol: PROSPERO CRD420251120318
- Data extraction forms: Available upon request
- Analysis code: Available upon request
- Raw extracted data: Available upon request

#### S10. Amendments to Protocol

No significant deviations from the registered protocol occurred.

#### S11. Author Contributions

Detailed author contributions:

- **Conceptualization**: A.G., M.O., E.K.
- **Search strategy development**: A.G., M.O.
- **Database searches**: A.G., M.O. (conducted independently and cross-verified)
- **Screening and selection**: A.G., M.O. (independent dual screening)
- **Data extraction**: A.G., M.O. (independent with consensus meetings)
- **Risk of bias assessment**: A.G., M.O., E.K. (E.K. served as arbitrator)
- **Statistical analysis**: A.G., M.O.
- **Manuscript drafting**: A.G. (first draft), M.O. (methods section)
- **Critical revision**: All authors (B.S.G., G.N.N. provided clinical expertise)
- **Supervision**: E.K., G.N.N. (co-senior authors)
- **Final approval**: All authors

#### S12. Protocol Registration and Amendments

- **PROSPERO Registration**: CRD420251120318
- **Registration Date**: [Prior to search initiation]
- **Protocol Amendments**: None
- **Deviations from Protocol**: None

#### S13. Search Updates

- **Initial Search**: August 5, 2025
- **Update Search**: Not performed (recent systematic review)
- **Alert Setup**: Bi-Monthly alerts established for ongoing monitoring

#### S14. Data Availability Statement

- **Extracted Data**: Available from corresponding author upon reasonable request
- **Analysis Code**: Available from corresponding author upon reasonable request
- **PRISMA Checklist**: Completed and available as supplementary file
- **Review Protocol**: Publicly available at PROSPERO (CRD420251120318)
